# Addressing Measurement Error of Machine-Learned Physical Activity in Nonlinear Dose-Response Survival Analysis: Development and Application of Accelerated Failure Time, Spline, and Simulation-Extrapolation Method

**DOI:** 10.64898/2026.08.25.26361155

**Authors:** Hiroshi Mamiya, Qihuang Zhang, Xiayi Zhang, Ying Yan, Abhinav Sharma

## Abstract

Wearable (accelerometer) data and machine-learning allow objective assessment of the amount of daily physical activity. However, wearable-derived human activity is subject to measurement error. No studies have corrected the dose-response association between physical activity and survival time to chronic diseases, including cardiovascular disease (CVD). The objective is to estimate the measurement error-corrected association between CVD events and multiple measures of daily duration of light and total physical activity, derived from machine-learning and conventional accelerometer-processing methods. Our method combined an accelerated failure time model, spline, and simulation-extrapolation (SIMEX). The method recovered the true dose-response non-linear association in simulated data, while the naive model failed to capture it due to substantial attenuation. Application to the UK Biobank accelerometer cohort also showed an increased protective association of total physical activity after SIMEX correction (Time Ratio [TR]=1.56, 95%CI:1.28–1.82 vs. TR=1.38, 95%CI:1.24–1.54 for SIMEX-corrected vs. uncorrected dose-response association between the 95th and 5th percentiles of total activity), with a similar increase for light physical activity. Sensitivity analysis indicates that the female population experiences a substantially larger protective association after SIMEX correction than than males. Dose-response survival analysis is a widely used analytical method in physical activity epidemiology and benefits from measurement error correction.

## Introduction

Insufficient Physical Activity (PA) is a major risk factor for Cardiovascular Disease (CVD) incidence and mortality, depression, cognitive decline, diabetes, and various cancers^1,2^. It is critical to characterize the dose-response association between CVD and the amount of PA, as it allows refining the PA guideline^3–5^. Dose-response meta-analysis of self-reported PA and incident CVD suggest a curvilinear or linear reduction of the incidence of CVD, typically estimated by a spline smoother combined with survival analysis^6^. Measurement error in self-reported PA is widely acknowledged, leading to attenuated associations between PA and health outcomes due to recall and social desirability bias^7^.

Accelerometer (wearable) data allow objective assessment of PA and are often used as a gold standard against self-report^2,7,8^. However, accelerometer-derived PA also exhibit measurement error, even when processed by the most recent activity signal detection methods such as machine learning (ML)^7–11^. The impact of measurement error in accelerometer-derived PA on etiologic association has been rarely investigated, and dose-response measurement error analysis has not been utilized even for self-reported PA. We have recently developed a method that combines spline and survival analysis as an Accelerated Failure Time (AFT) model, with measurement error correction via Simulation-Extrapolation (SIMEX) hereafter called AFT-Spline-SIMEX^12–14^.

Our objective to estimate the measurement error-corrected dose-response association between wearable-derived duration of PA and survival time to CVD using AFT-Spline-SIMEX. We will compare these estimates with *naive* method without error-correction using simulated CVD and PA data, followed by empirical investigation using the UK Biobank accelerometer sub-study.

The study includes two intensity-based PA exposures: the daily duration of Light PA (LPA) and Total PA (TPA, the combination of LPA and Moderate-Vigorous PA). Moderate-Vigorous PA is also a commonly used intensity-based exposure definition; however, we did not use it in this study, as it may require a separate approach to address zero-inflation (i.e., many participants in the validation data have zero minutes of Moderate-Vigorous PA). LPA and TPA are critical exposures for PA guidelines, based on findings suggesting that accumulating any or low-intensity of PA is critical^2,15,16^.

Each exposure intensity was represented by three PA measures derived from three accelerometer processing methods: (1) Low-pass Filtered Euclidean Norm Minus One (LFENMO), (2) ML, and (3) an activity counts algorithm^17,18^. These methods represent the diverse, non-standardized approaches currently used to process accelerometer data, which can yield substantially different amounts of PA from the same data^11^. Rather than relying on a single derived measure, our AFT-Spline-SIMEX framework integrates multiple disparate measures into a unified model.

## Methods

### Study Design

This was a longitudinal (time-to-event) study using simulation and empirical analysis using UK Biobank between 2013 and 2022 inclusive. We first describe the UK Biobank data, including validation data, followed by simulated data and the statistical method.

### Data and population

UK Biobank is a prospective cohort consisting of approximately 500,000 participants aged 40–69 at baseline in 2006^19^. Our study population is the UK Biobank accelerometer sub-cohort containing 103,686 participants wearing wrist-worn Ax3 devices (Axivity, United Kingdom) for seven consecutive days between 2013 and 2014^18^. Individuals with insufficient wear time (less than 72 h)^18^, those who developed incident CVD before accelerometer data collection, and those lost to follow-up were excluded (UK Biobank removes entire follow-up records among those who withdrew). We also removed those missing baseline covariates, e.g., diets, income, education, and smoking, which were less than 5% of participants, leading to a sample size of 90,237. Supplementary Figure S3 shows the resulting exclusion flowchart.

#### Three exposure measures for TPA and LPA

UK Biobank calibrated raw tri-axial accelerometer to local gravity, imputed non-wear time, and computed a summary vector magnitude^18^. The resulting five-second summary of acceleration minus one gravitational unit is the first exposure measure, LFENMO^20^. LFENMO provided the daily duration spent on TPA and LPA by applying a validated intensity cut-point of between 50-100 mg for LPA and TPA as the duration of any intensity exceeding the 50mg cut-point^21,22^. UK Biobank also provides pre-computed daily duration of LPA and TPA by ML, which is the combination of random forest and hidden Markov model^23^. For the third measure (activity counts), we applied an open-source method to reproduce a previously proprietary activity counts metric from the processed tri-axial accelerometer data described above^24,25^. We then computed the durations of TPA and LPA using validated intensity cut-points for wrist-worn devices ^26^. These are 2860–3940 counts per minute (CPM) for LPA and ≥ 3941 CPM cut-points for Moderate-Vigorous PA, multiplied by 0.5 for our 30-second time window. While not used in this study, the algorithm to exactly reproduce activity counts from raw accelerometer data has been available since 2022^27^.

#### Outcome

The outcome is the first episode of stroke or myocardial infarction (both fatal and non-fatal) as ascertained by the UK Biobank’s adjudicated algorithms integrating hospital episodes, death certificates, and self-reports^28^. Follow-up of participants began after the participant-specific date of accelerometer data collection and continued until death, incident CVD, or the last available date of the outcome records, on November 30, 2022.

#### Validation data

We used external validation data that contain both ground-truth PA and device-derived PA measures. The validation study, *Capture-24*, comprise 151 participants who wore a portable camera for 24 hours in Oxfordshire, UK, concurrently collected with the accelerometer sub-cohort data in the UK Biobank using the same accelerometer device (wrist-worn Ax3)^29^. To our knowledge, this is the largest validation dataset for free-living hour activity, involving manual annotation of camera images at 30-second intervals using a validated protocol^29^. The durations of LPA and TPA were computed by aggregating the annotated images as ground truth. The accelerometer data were processed by the identical methods used in the UK Biobank data to generate the three derived measures.

### Simulation

We conducted a simulation (*R* = 500 replicates of *n* = 2000 participants) to assess whether the AFT–spline–SIMEX estimator recovers a nonlinear dose-response association under measurement error. Event times were generated from an AFT model in which the true dose-response curve rises from a threshold at 150 minutes/day of LPA and plateau at approximately 300 minutes/day^30^, with approximately 76% of participants experiencing the event. Three error-contaminated exposure measures of LPA were then generated with correlated classical additive errors on the log scale, scaled to a mean per-measure reliability of 0.50, together with an external validation sample used to estimate the error covariance. The data-generating process, the estimators compared, and the performance metrics are detailed in Supplementary Appendix Supplementary Appendix S2.7.

### Statistical method

Our AFT-spline-SIMEX estimator uses a natural cubic spline on the exposure and corrects for measurement error in multiple derived measures per exposure in two-steps. In Step 1, it captures the error structure using validation data and combines the three PA measures into *a single calibrated exposure measure*. In Step 2, it applies SIMEX to obtain bias-corrected estimates of the non-linear exposure–outcome association using the combined exposure.

#### Step 1: Measurement error characterization

The three derived durations of PA exhibit measurement error, which we modeled on the log scale with normally distributed errors. These are LFENMO 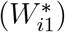, ML 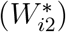, and activity counts 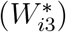. Let *W*_*i*_ denote the (unobserved) true daily LPA for participant *i*, and let 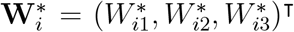 denote the vector of error-contaminated durations. We specified a multivariate log-linear measurement error model,

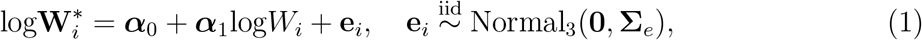

where ***α***_0_ = (*α*_01_, *α*_02_, *α*_03_)^⊤^ and ***α***_1_ = (*α*_11_, *α*_12_, *α*_13_)^⊤^ are three measure-specific intercepts and slopes. A 3 × 3 residual covariance matrix is denoted as **Σ**_*e*_, whose off-diagonal entries quantify the correlation between measurement errors of the three measures sharing the same underlying accelerometer signal. Of note, Model (1) is a linear (calibration) measurement error model, as ***α***_1_ is estimated instead of being fixed at one often considered in the classic model. The parameters 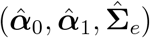 were estimated from the Capture-24 validation study.

We combined the three derived measures using generalized least squares (GLS). Each measure was first back-transformed onto the scale of log*W*_*i*_ as 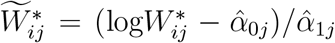, such that 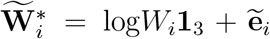, where the back-transformed error 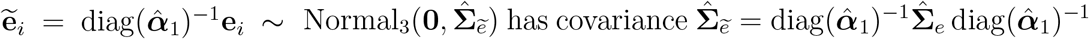. The GLS-combined exposure is given by

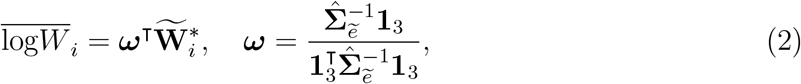

with 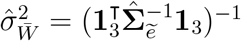 being the residual variance. The combiner ***ω*** is the best linear unbiased combination of the three calibrated measures and reduces to inverse-variance weighting when 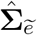 is diagonal. The back-transformation returns the combined exposure to the classical addit ive form, 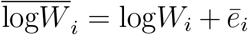 with 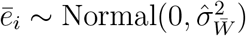, which is the error structure required by SIMEX.

#### Accelerated failure time model with spline and confounders

Let *T*_*i*_ denote the time to the outcome (i.e, CVD) for participant *i*, and let **Z**_*i*_ denote the vector of confounders. We specified an AFT model of the form

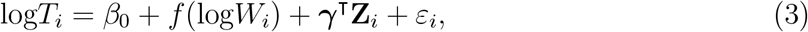

where *f* (·) is a natural cubic spline with one interior knot at the 50th percentile of the observed PA distribution to capture curvilinear association with smooth and monotonically diminishing, i.e., decelerating protective effects^30^. A specification with two interior knots placed at the 33rd and 67th percentiles was also tested to capture a more flexible shape. We also considered ***γ***, a vector of confounder coefficients, and *ε*_*i*_ follows an extreme-value distribution, so that *T*_*i*_ is Weibull-distributed. The log-normal alternative is also considered and compared by Akaike’s Information Criterion (AIC) in the sensitivity analysis. The exposure log*W*_*i*_ in Model (3) can be the combined GLS exposure, 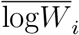, from (2) or one of the three exposure measures. This is our naive model, which is biased when the exposure is measured with error^31–33^, to be compared with the SIMEX-corrected version of this model in Step 2 below.

We ran four naive and four SIMEX models treating the exposure as: 1) GLS-combined measure, 2) LFENMO only, 3) ML only, and 4) activity counts only. The associations from these models were compared against the corresponding SIMEX-corrected ones. The process was repeated for the LPA and TPA exposures, respectively, resulting in 16 models in total.

Confounders are age (continuous), sex (binary), and the UK Biobank-defined ordered categories for education, income, and the consumption of tobacco smoking, alcohol, red meat, fruits, vegetables, and oily and non-oily fish. We also controlled for ethnic background, dichotomized into visible minorities (Asian or Asian British, Black or Black British, and other non-White groups) vs. White, given the limited proportion of each non-White group. We did not include Body mass index, blood cholesterol, and medication, since they were potential mediators of PA and the outcome, and their addition did not change the dose-response association in our and others’ previous studies using the same data^11,34^ .

#### Step2: SIMEX correction

SIMEX is a widely used method to correct measurement error in epidemiology^35^. Briefly, it inflates the error variance in the observed data, and the resulting bias in parameter estimates is extrapolated back to the error-free case^12,14^. For a grid of error inflation factors *λ* ∈ {0.1, 0.2,…, 2.0} (20 equally spaced values), we simulated *B* = 20 error-contaminated datasets with the exposure. The AFT model (3) was fitted to each error-added dataset, and the fitted dose-response curve was averaged across the *B* replicates at each *λ*. A dense inflation grid with few replicates per grid point is sufficient for stable extrapolation^36^. At each point of the exposure grid, the averaged estimates 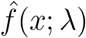 were then modelled as a quadratic function of the inflation factor and extrapolated to *λ* = −1, the hypothetical case of no measurement error, giving the SIMEX-corrected dose-response curve 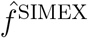 across the range of 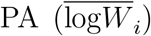. Extrapolating the curve pointwise, rather than extrapolating the spline coefficients, keeps the correction independent of the basis used to represent the curve at each *λ*.

#### Variance Estimation

We used a novel two-stage nonparametric bootstrap to propagate sampling uncertainty from the Step 1 calibration (Capture-24 validation study, *n* = 151) and the Step 2 main analysis. The approach provides valid (wider) confidence intervals for the SIMEX-corrected dose-response curve to avoid false positives. Specifically, on each bootstrap replicates, we (i) drew a sample of the validation participants and refit the measurement error model (1) to obtain bootstrap replicate-specific calibration estimates 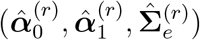; (ii) recomputed the GLS weights ***ω***^(*r*)^ and the conditional residual variance 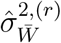 from these refit estimates; (iii) drew a nested bootstrap sample of the study sample (i.e., UK Biobank) and combined (2) ; and (iv) ran the SIMEX correction with assumed error variance 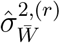 to obtain the corrected dose-response curve 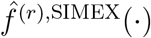. This nested schema avoids the optimistic (falsely narrow) 95% CI that would result from erroneously treating the calibration estimates as fixed^37,38^.

#### Sensitivity analyses

We replaced the Weibull distribution in our AFT model with another commonly used distribution: the lognormal distribution. We also performed sex-specific analysis of dose-response association due to known sex-specific heterogeneity in CVD etiology^39^. In addition, we used an alternative volume-based definition of TPA, in which each minute of moderate-to-vigorous PA was counted as two minutes to reflect its potentially greater protective effects^40^. Fourth, we used a subset of validation data (71 out of 151 participants) that had an overlapping age range with UK Biobank: participants in the validation study were younger, with an approximate median age of 39.9 based on the categorical age range in the validation data vs 66.2 in the UK Biobank accelerometer sub-cohort).

Analyses were conducted in R (version 4.4.1; R Foundation for Statistical Computing, Vienna, Austria). We obtained approval from the Faculty of Medicine Institutional Review Board (A08-M55-24A). We provided the Strengthening the Reporting of Observational studies in Epidemiology checklist (Supplementary File 3).

## Results

### Simulated data

Figure 1 shows the simulation results. Ignoring measurement error produced a substantially attenuated curve (panel B), whereas the SIMEX-corrected estimate (panel C) recovered the true dose-response shape (panel A). The two-stage bootstrap recovered the sampling variability of the corrected curve (standard error ratio 1.05; Appendix Supplementary Appendix S2.7.5). A conventional one-stage bootstrap, which treats the calibration as known, gives intervals that are too narrow: with a small validation study the two-stage interval was considerably wider (Figure 2), and the difference narrows as the validation study grows (Table 1 and Appendix Supplementary Appendix S2.7.6).

**Table 1.** Two-stage versus one-stage bootstrap as the validation sample shrinks relative to the main study (*n* = 2000). Coverage is the pointwise empirical coverage of the 95% interval over the interior 5%–95% region; width inflation is the ratio of mean interior interval widths (two-stage over one-stage).

| $n_{\text{val}}$ | ratio $n/n_{\text{val}}$ | Two-stage cov. | One-stage cov. | Width inflation |
| --- | --- | --- | --- | --- |
| 500 | 4 | 0.864 | 0.848 | 1.04 |
| 150 | 13 | 0.839 | 0.815 | 1.07 |
| 50 | 40 | 0.898 | 0.826 | 1.25 |

Entries are based on $N = 50$ outer replicates with $R = 120$ inner bootstrap resamples. The $n_{\text{val}} = 500$ row corresponds to the data-generating process used throughout Appendix Supplementary Appendix S2.7.1; the two-stage coverage reported for that process in Appendix Supplementary Appendix S2.7.5 (85.0%) comes from a separate run with $N = 100$ outer replicates, and the two figures agree within Monte Carlo error.

**Figure 1.**
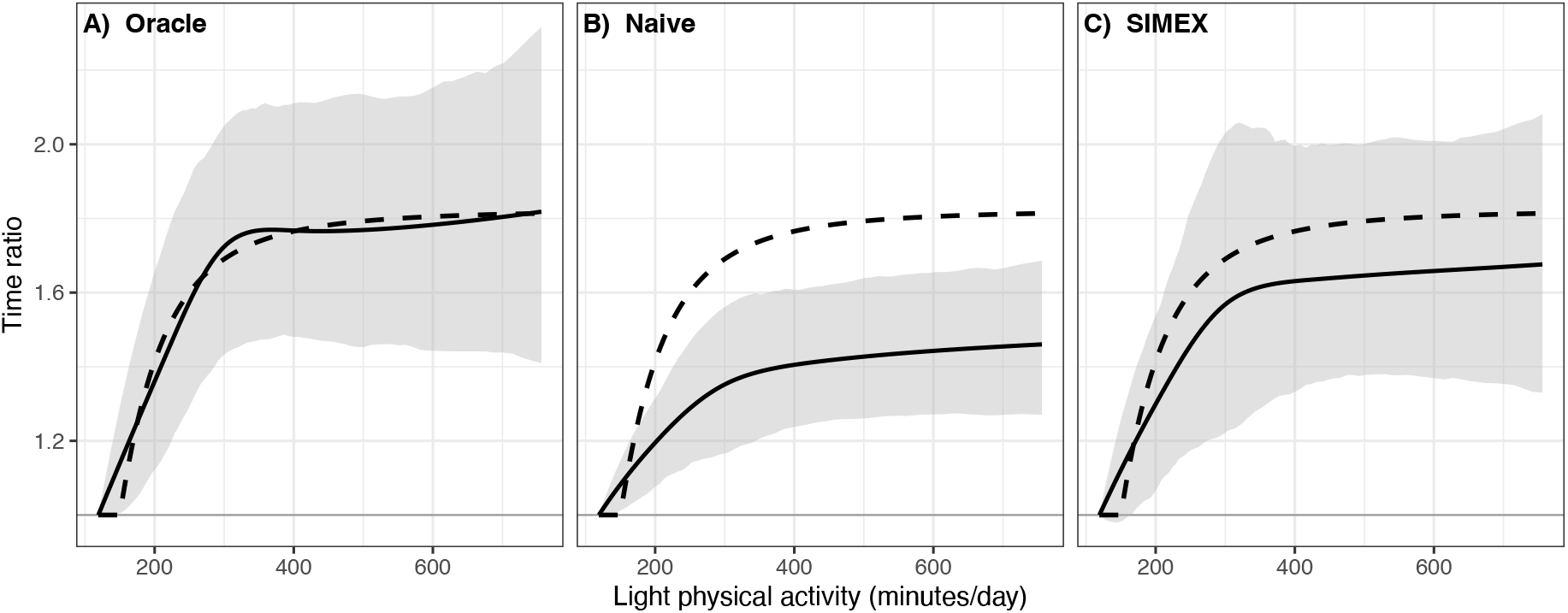
Estimated dose-response association on simulated daily light physical activity in minutes. In each panel, the solid line is the mean time-ratio curve across *R* = 500 Monte Carlo replicates. The shaded band is the pointwise 2.5%–97.5% empirical envelope. **(A)** Oracle; **(B)** error-uncorrected pooled Generalized Least Square-estimated measure from three exposure measures, which fails to capture the true dose-response function due to attenuation; and **(C)** SIMEX-corrected calibrated measure, which recovers the true dose-response curve.

**Figure 2.**
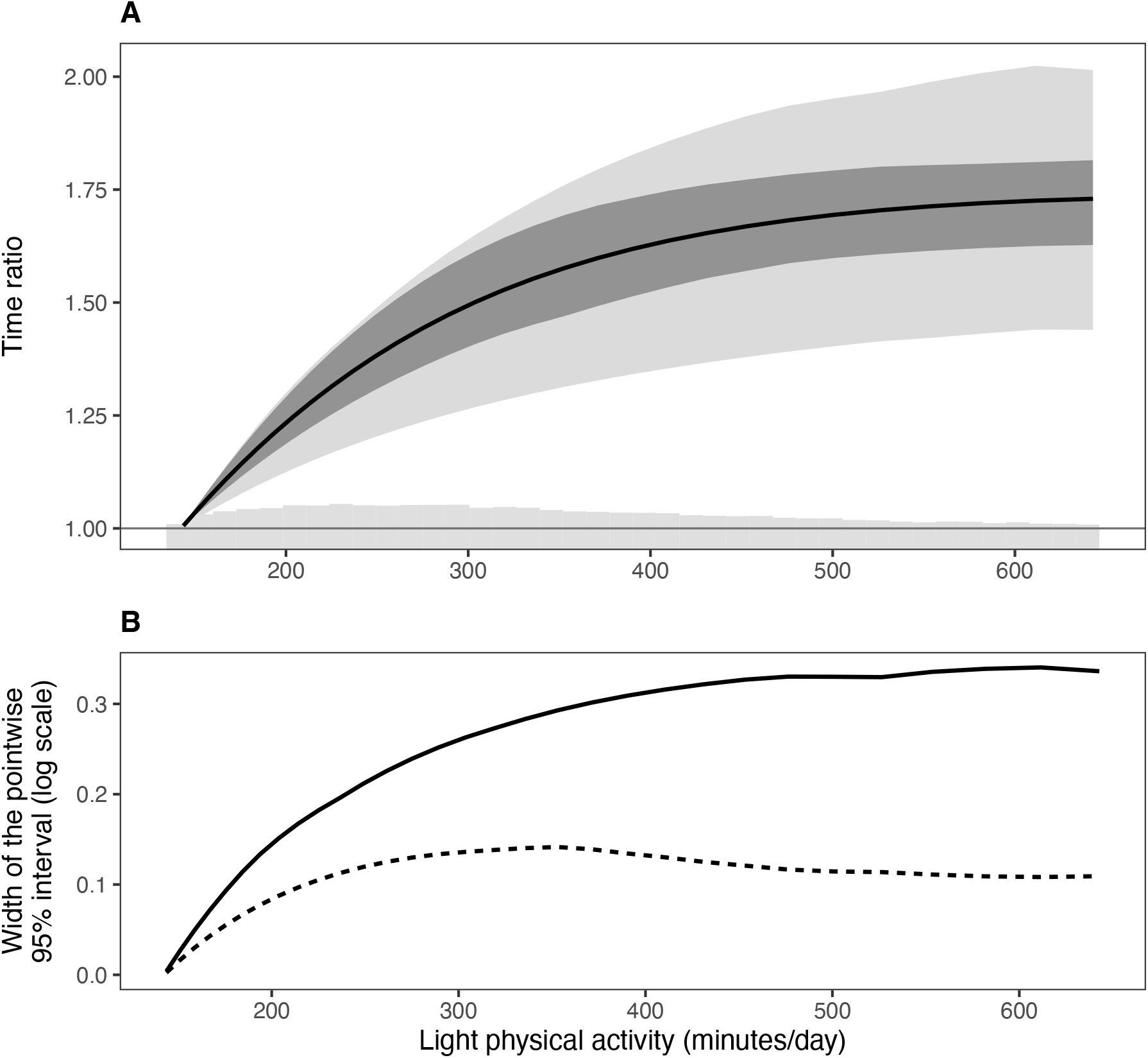
What the validation-aware two-stage bootstrap costs in interval width when the validation sample is small (simulated data, *n* = 15,000, *n*_val_ = 25, *R* = 80). The validation sample here is deliberately smaller than Capture-24 (*n*_val_ = 151) so that the contrast between the two designs is visible; Table 1 reports the width inflation across validation sizes spanning that of the present study. **(A)** SIMEX-corrected dose-response curve with pointwise 95% intervals from both designs: the *outer, lighter* band is the two-stage interval, which resamples the validation study in addition to the main study, and the *inner, darker* band is the one-stage interval, which holds the Step 1 calibration fixed. The two designs differ only in how the bootstrap replicates are drawn, not in the estimate itself; the single black curve is the two-stage point estimate. The grey histogram at the base shows the exposure distribution. **(B)** Width of the same two intervals across the exposure: the *solid* line is the two-stage interval and the *dashed* line the one-stage. Averaged over the range shown, the two-stage interval is 2.2 times wider. Both panels are restricted to the interpretable region between the 5th and 95th percentiles of the exposure, since interval widths compared in the extrapolation region would not be meaningful.

### Validation data

The magnitude of error relative to ground truth varied widely across the three derived measures for both LPA (Figure 3) and TPA (Figure 4). ML showed the highest agreement and activity counts showed the lowest agreement. Activity counts and LFENMO also underestimated the durations of LPA and TPA relative to ground truth (Supplementary Figures S1 and S2)

**Figure 3.**
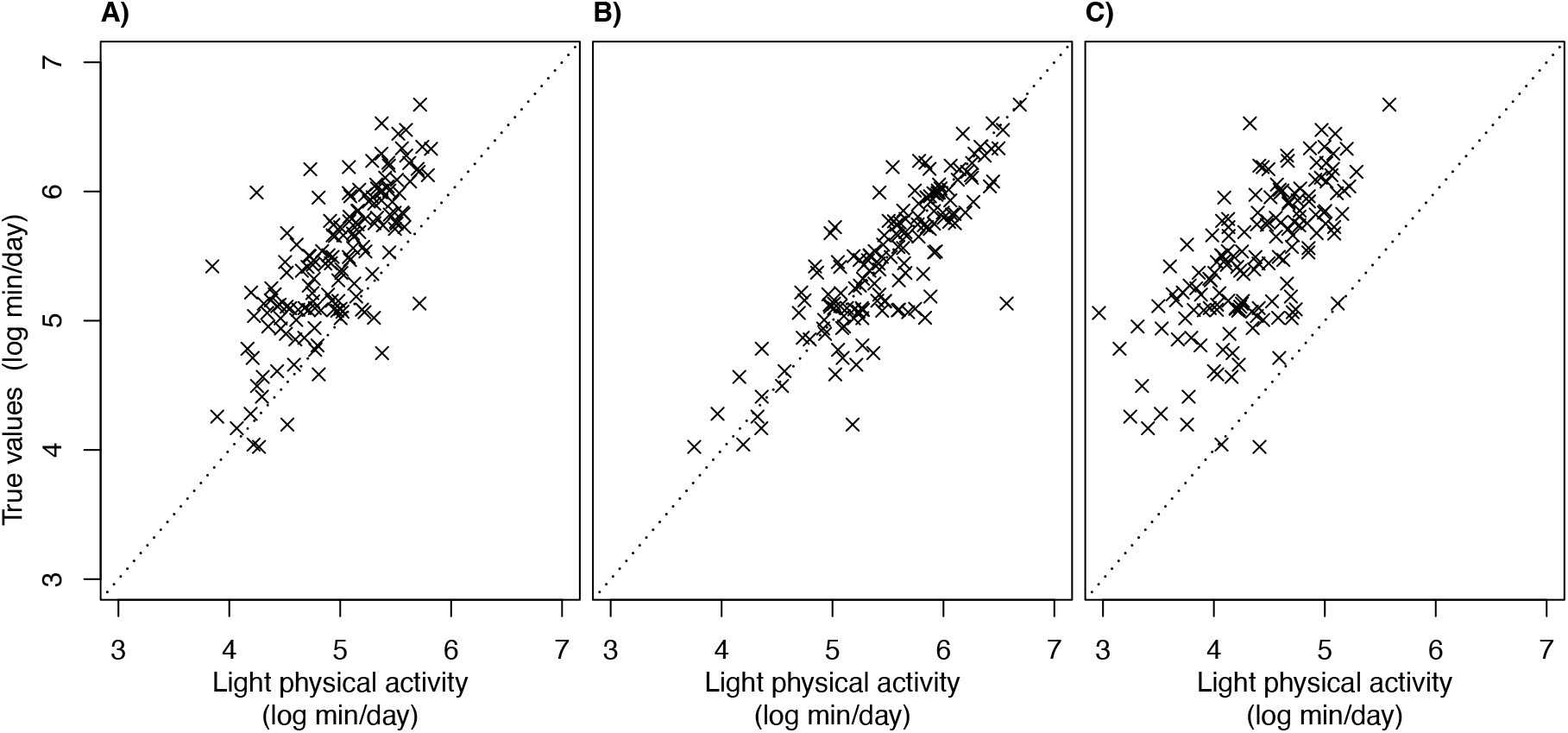
Relationship between log-transformed light physical activity duration measured by ground truth and accelerometer-derived measures: **(A)** low-pass filter Euclidean Norm Minus One, **(B)** machine learning, and **(C)** activity counts.

**Figure 4.**
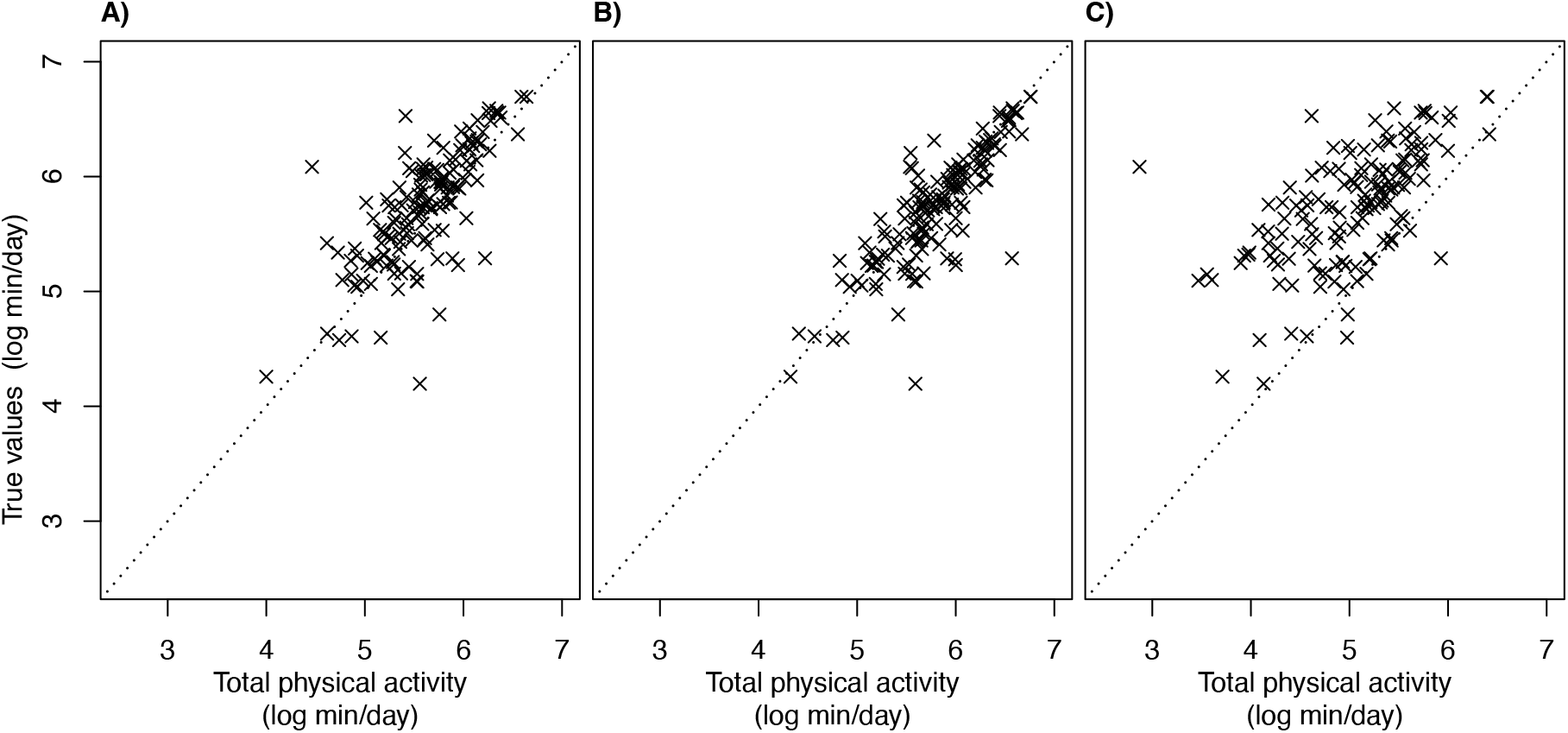
Relationship between log-transformed total physical activity duration measured by ground truth and accelerometer-derived measures: **(A)** low-pass filter Euclidean Norm Minus One, **(B)** machine learning, and **(C)** activity counts.

### UK Biobank accelerometer sub-study

Descriptive analysis indicates that the derived durations of LPA and TPA in UK Biobank were much larger for ML than LFENMO and activity counts (Table 2). Also, all derived durations are larger among participants with incident CVD than those without CVD.

**Table 2.** Distribution of participant characteristics by outcome status among adults in the UK Biobank accelerometer sub-cohort, 2013–2022.

| Baseline Characteristics | Cardiovascular disease<br>N = 2,841 | No cardiovascular<br>disease<br>N = 86,989 |
| --- | --- | --- |
| Follow-up time in months, Median (25th, 75th) | 52.4 (29.2, 75.4) | 96.9 (90.3, 103.0) |
| Light physical activity: Low-pass Filter Euclidean Norm Minus One, Median (25th, 75th) | 140.0 (113.8, 167.6) | 145.5 (121.6, 171.6) |
| Light physical activity: Machine learning, Median (25th, 75th) | 281.9 (216.2, 351.0) | 297.4 (235.7, 366.1) |
| Light physical activity: Activity count, Median (25th, 75th) | 84.3 (65.2, 106.3) | 90.0 (71.0, 110.6) |
| Total physical activity: Low-pass Filter Euclidean Norm Minus One, Median (25th, 75th) | 198.3 (157.5, 241.7) | 216.3 (176.4, 259.2) |
| Total physical activity: Machine learning, Median (25th, 75th) | 318.5 (249.5, 396.8) | 341.3 (274.6, 412.5) |
| Total physical activity: Activity count, Median (25th, 75th) | 152.7 (109.5, 204.0) | 178.1 (131.3, 230.5) |
| Age, Median (25th, 75th) | 67.7 (62.8, 71.5) | 63.0 (55.9, 68.3) |
| Ethnicity, n (%) |  |  |
| Non-white | 79 (2.8) | 2,594 (3.0) |
| White | 2,762 (97.2) | 84,395 (97.0) |
| Sex, n (%) |  |  |
| Female | 1,092 (38.4) | 50,485 (58.0) |
| Male | 1,749 (61.6) | 36,504 (42.0) |
| Education, n (%) |  |  |
| None of the below | 381 (13.4) | 6,681 (7.7) |
| O levels/GCSEs or equivalent, CSEs or equivalent | 698 (24.6) | 21,315 (24.5) |
| A levels/AS, NVQ/HND/HNC or equivalent | 724 (25.5) | 20,486 (23.6) |
| College or University degree | 1,038 (36.5) | 38,507 (44.3) |
| Household income in British Pounds, n (%) |  |  |
| Less than 18,000 | 562 (19.8) | 10,948 (12.6) |
| 18,000 to 30,999 | 709 (25.0) | 18,831 (21.6) |
| 31,000 to 51,999 | 724 (25.5) | 22,832 (26.2) |
| 52,000 to 100,000 | 460 (16.2) | 20,374 (23.4) |
| Greater than 100,000 | 96 (3.4) | 5,955 (6.8) |
| Do not know/Prefer not to answer | 290 (10.2) | 8,049 (9.3) |
| Smoking, n (%) |  |  |
| Never | 1,356 (47.7) | 50,544 (58.1) |
| Previously | 1,224 (43.1) | 30,675 (35.3) |
| Currently | 261 (9.2) | 5,770 (6.6) |
| Alcohol consumption, n (%) |  |  |
| Never | 203 (7.1) | 4,728 (5.4) |
| Less than once a week | 574 (20.2) | 17,593 (20.2) |
| Once or twice a week | 674 (23.7) | 21,919 (25.2) |

| Baseline Characteristics | Cardiovascular disease<br>N = 2,841 | No cardiovascular disease<br>N = 86,989 |
| --- | --- | --- |
| Three or four times a week | 698 (24.6) | 22,830 (26.2) |
| Daily or almost daily | 692 (24.4) | 19,919 (22.9) |
| Processed meat, n (%) |  |  |
| Less than 2 times a week | 1,903 (67.0) | 62,097 (71.4) |
| 2–4 times a week | 822 (28.9) | 21,832 (25.1) |
| More than 4 times a week | 116 (4.1) | 3,060 (3.5) |
| Red meat, n (%) |  |  |
| Less than 2 times a week | 394 (13.9) | 15,847 (18.2) |
| 2–4 times a week | 983 (34.6) | 29,458 (33.9) |
| More than 4 times a week | 1,464 (51.5) | 41,684 (47.9) |
| Oily fish, n (%) |  |  |
| Less than 2 times a week | 2,247 (79.1) | 71,962 (82.7) |
| 2–4 times a week | 567 (20.0) | 14,336 (16.5) |
| More than 4 times a week | 27 (1.0) | 691 (0.8) |
| Non-oily fish, n (%) |  |  |
| Less than 2 times a week | 2,389 (84.1) | 73,810 (84.8) |
| 2–4 times a week | 443 (15.6) | 12,767 (14.7) |
| More than 4 times a week | 9 (0.3) | 412 (0.5) |
| Fresh fruit, n (%) |  |  |
| Less than 2 servings a day | 1,007 (35.4) | 28,299 (32.5) |
| Between 2 and 4 servings a day | 1,638 (57.7) | 52,183 (60.0) |
| More than 4 servings a day | 196 (6.9) | 6,507 (7.5) |
| Cooked vegetables, n (%) |  |  |
| Less than 2 servings a day | 437 (15.4) | 14,848 (17.1) |
| Between 2 and 4 servings a day | 2,115 (74.4) | 64,069 (73.7) |
| More than 4 servings a day | 289 (10.2) | 8,072 (9.3) |

*Note:* 25th indicates the first quartile and 75th indicates the third quartile.

For the dose-response association, the combined GLS and ML showed nearly identical shapes: a weakly curvilinear increase in survival time as the daily duration of LPA accumulates (Figure 5, Panels A and C, respectively). This suggests that ML drove the shape of the GLS-combined exposure. LFENMO showed a strong curvilinear protective association with a distinct plateau for both SIMEX-corrected and naive estimates (panel B); however, the plateaued region was located nearly outside the interpretable region (inner 90 percent of LPA distribution enclosed by vertical dotted lines). SIMEX-corrected association showed a stronger protective association for ML, LFENMO, and GLS than naive analysis. In contrast, for activity counts, SIMEX-corrected association was nearly identical to that of naive association, with a far narrower distribution of TPA than other measures.

**Figure 5.**
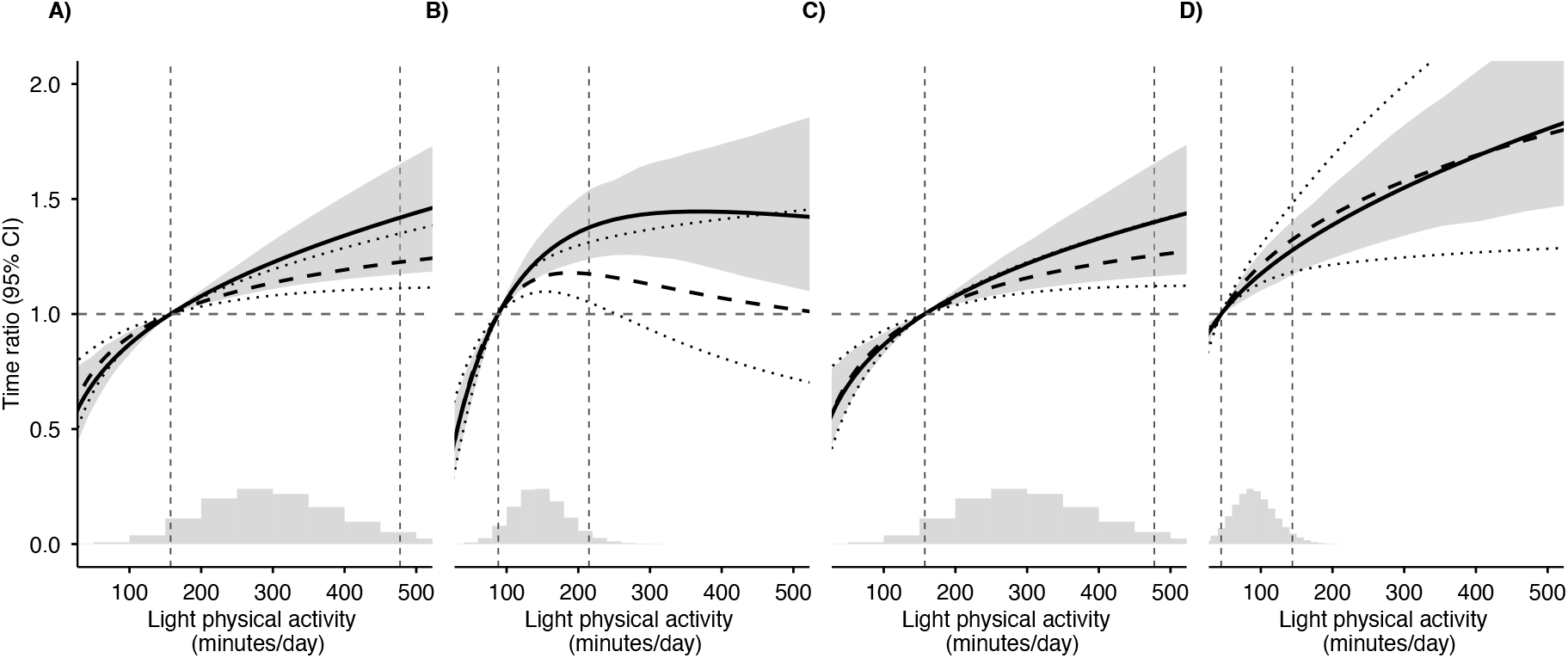
Comparison of dose-response associations between light physical activity and cardiovascular outcome for four exposure measures fit in four models: **(A)** Generalized Least Square Combined exposure; **(B)** low-pass filter Euclidean Norm Minus One; **(C)** machine learning; and **(D)** activity counts. SIMEX-corrected curves are indicated by solid black line with a grey band for 95% CI. Naive estimates are indicated by dashed line, with dotted 95% CI. Observed-exposure distribution is indicated by a gray histogram at the bottom. The region outside the 5th and 95th percentile of the exposure distribution, as indicated by dashed vertical lines, is not supported by the observed data and thus should not be interpreted. Horizontal dashed line indicates a time ratio of 1.00 as the null association, referenced at the 5th percentile of each exposure.

Table 3 presents the ratio of survival time comparing the 95th and 5th percentiles of LPA. Because accelerometer processing methods estimate substantially different durations of LPA, the absolute durations corresponding to these percentiles vary across methods (vertical dashed lines in Figures 5 and 6); hence, we use percentile-based comparison. Supporting Figure 5, SIMEX-correction led to a larger time ratio (stronger protective effects) for the GLS-combined exposure (Time Ratio [TR] = 1.42; 95%CI:1.17–1.65) than the naive estimate (TR=1.23; 95%CI:1.11-1.35). The 95%CI after SIMEX correction was narrower for activity counts but similar for the other measures. In contrast, activity counts-derived LPA showed early identical time-ratio (TR=1.28, 95%CI; 1.17-1.40 after SIMEX correction, TR=1.32, 95%CI;1.18-1.48).

**Table 3.** Increased survival time as light physical activity increases from the 5th percentile to the 95th percentile for SIMEX-corrected model and naive model, for each of four exposure measures fit in four models. Percentiles are specific to each exposure measure, so the contrasts span different ranges of daily minutes and are not directly comparable across measures.

| Exposure measure | Light physical activity (min/day) |  | Time ratio |  |
| --- | --- | --- | --- | --- |
|  | 5th %tile (reference) | 95th %tile | SIMEX | Naive |
| GLS (combined) | 157 | 477 | 1.42 (1.17–1.65) | 1.23 (1.11–1.35) |
| LFENMO | 88 | 215 | 1.38 (1.24–1.54) | 1.17 (1.05–1.31) |
| Machine Learning | 157 | 477 | 1.40 (1.16–1.65) | 1.25 (1.12–1.40) |
| Activity count | 45 | 144 | 1.28 (1.17–1.40) | 1.32 (1.18–1.48) |

*Abbreviations:* GLS, calibrated generalized least-squares combiner; LFENMO, low-pass filter Euclidean norm minus one; SIMEX, simulation–extrapolation; %tile, percentile.

**Figure 6.**
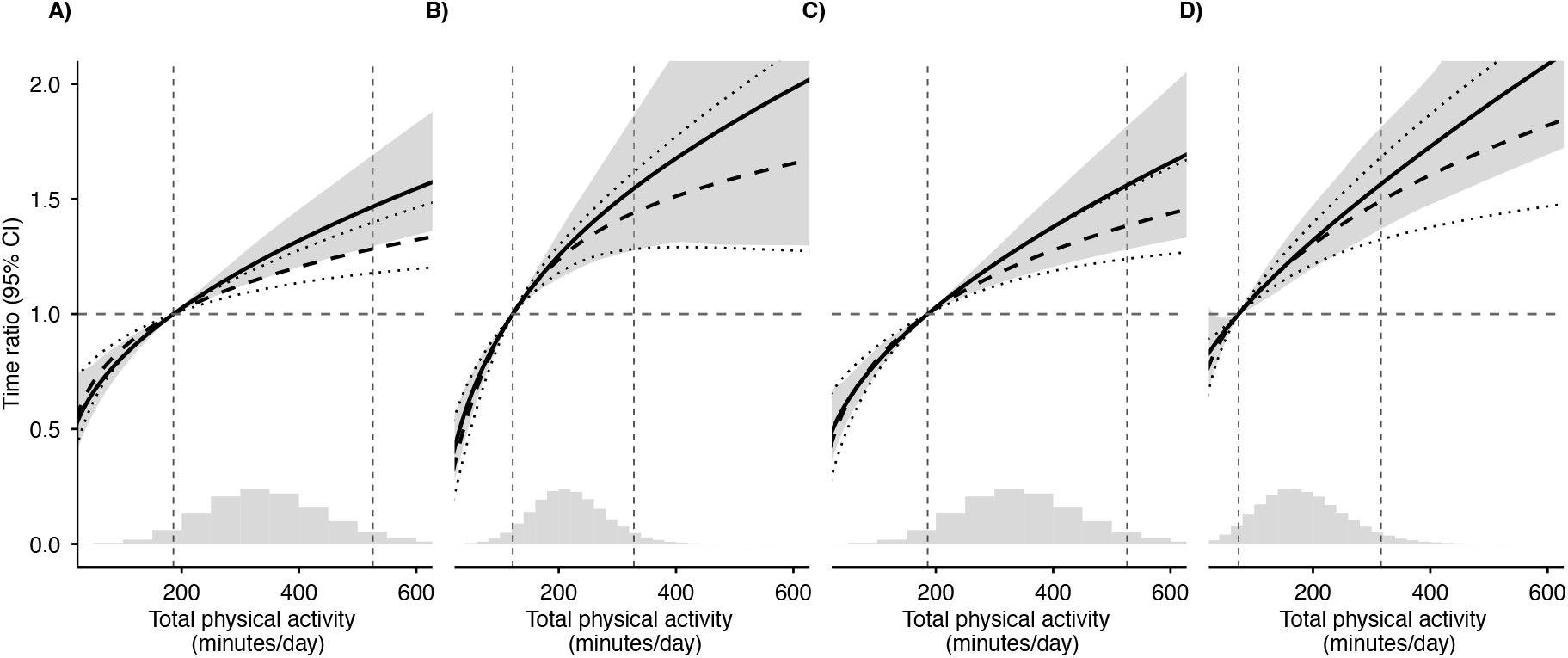
Comparison of dose-response associations between total physical activity and cardiovascular outcome for four exposure measures fit in four models:: **(A)** Generalized Least Square Combined exposure; **(B)** low-pass filter Euclidean Norm Minus One; **(C)** machine learning; and **(D)** activity counts. SIMEX-corrected curves are indicated by solid black line with a grey band for 95% CI. Naive estimates are indicated by dashed line, with dotted 95% CI. Observed-exposure distribution is indicated by a gray histogram at the bottom. The region outside the 5th and 95th percentile of the exposure distribution, as indicated by dashed vertical lines, is not supported by the observed data and thus should not be interpreted. Horizontal dashed line indicates a time ratio of 1.00 as the null association, referenced at the 5th percentile of each exposure.

TPA also showed a weakly curvilinear dose-response shape (Figure 6), with a stronger (steeper) association than LPA. As in LPA, SIMEX correction further enhanced the protective associations for the GLS-combined exposure and ML (panels A and C, respectively), but showed a smaller difference for LFENMO and little difference for activity counts (panels B and D, respectively). The 95%CI was wider for ENMO and ML after SIMEX correction.

As in LPA, the comparison of time ratios shows that SIMEX correction led to a greater gain in survival time than the naive estimates for all four TPA surrogates (Table 4). The gain after SIMEX correction was the strongest for the GLS-combined exposure (TR=1.47; 95%CI,1.30-1.69 for SIMEX, and TR = 1.28; 95%CI, 1.18-1.40 for naive). In contrast, activity counts showed little increase of TA (TR=1.56, 95%CI; 1.37-1.81 after SIMEX correction, TR=1.49, 95%CI;1.32-1.68).

**Table 4.** Increased survival time as total physical activity increases from the 5th percentile to the 95th percentile for SIMEX-corrected model and naive model, for each of four exposure measures fit in four models. Percentiles are specific to each exposure measure, so the contrasts span different ranges of daily minutes and are not directly comparable across measures.

| Exposure measure | Total physical activity (min/day) |  | Time ratio |  |
| --- | --- | --- | --- | --- |
|  | 5th %tile (reference) | 95th %tile | SIMEX | Naive |
| GLS (combined) | 186 | 526 | 1.47 (1.30–1.69) | 1.28 (1.18–1.40) |
| LFENMO | 122 | 328 | 1.55 (1.28–1.86) | 1.44 (1.28–1.62) |
| Machine Learning | 186 | 526 | 1.56 (1.28–1.82) | 1.38 (1.24–1.54) |
| Activity count | 73 | 316 | 1.56 (1.37–1.81) | 1.49 (1.32–1.68) |

*Abbreviations:* GLS, calibrated generalized least-squares combiner; LFENMO, low-pass filter Euclidean norm minus one; SIMEX, simulation–extrapolation; %tile, percentile.

The results above are based on spline with a single internal knot. Dose-response associations based on two internal knots showed a more distinct curvilinear association, with LFENMO showing a more prominent protective association after SIMEX correction for LPA (Supplementary Figure S4); the corresponding results for TPA are shown in Supplementary Figure S5. The two specifications of spline (one vs two internal knots) showed similar fit (ΔAIC *<* 4). However, the results for the GLS-combined exposure and ML showed a slight decrease in association, i.e., an inverse U-shape, which is inconsistent with the previously reported monotonically increasing association for LPA and TPA and thus not discussed further.

### Sensitivity analysis

Use of a lognormal distribution resulted in higher values of AIC than fits based on Weibull for all models, thus excluded from analysis. Sex-specific analysis shows a stronger protective association among females than males for both LPA and TPA (Figures S6 and S7, respectively). More importantly, SIMEX appears to increase the steepness of association only among females. The use of the volume-based TPA weighted by Moderate-Vigorous PA showed a more prominent increase than the unweighted TPA (Supplementary Figure S8). Finally, the use of a subset of the validation restricted to the age range of the UK Biobank showed a much stronger association for LFENMO and a slightly stronger association for activity counts with much wider 95%CI, but did not show any changes from the main analysis for ML and the GLS-combined measure (Supplementary Figures S10 and S9).

## Discussion

Wearable-derived PA is subject to measurement error, even with the growing use of ML to quantify PA from accelerometer data. We used a novel measurement error model for dose-response survival analysis, with a focus on CVD. Our AFT-Spline-SIMEX model correctly recovered the true association in simulated data, whereas naive analysis showed attenuation. An empirical application to the UK Biobank accelerometer sub-corhot confirmed the simulation results: the stronger protective association of PA derived from the combined exposure, ML, and LFENMO after accounting for measurement error. However, the association derived from activity counts changed little after SIMEX correction.

The larger measurement errors for LFENMO and activity counts than for ML in the validation data are expected. This is because they typically use intensity cut-points calibrated and validated in an external population, often from participants performing programmed exercise in a laboratory rather than in free-living conditions. ML is reported to be more accurate than cut-point methods, since it is trained on a large number of predictive features from accelerometer data, enabling more personalized PA intensity predictions. ML’s superior performance in this study also rests on the model being trained on the Capture-24 validation data, which is collected from free-living participants.

Not surprisingly, ML drove the shape of the protective dose-response association from the combined GLS exposure measure due to the much smaller error variance relative to the ground truth, leading to large inverse error weights upon pooling the three PA measures. The overall shapes of dose-response associations from all exposure measures agree with previous dose-response meta-analyses: the sharpest increase in protective association at lower PA durations, regardless of exposure measures, with a slightly decelerated or diminished (plateaued) association as PA accumulates. SIMEX correction further increased these protective associations for both intensities (LPA and TPA). Thus, existing naive analyses and dose-response meta-analyses may have reported attenuated benefits of LPT and TPA, except possibly for activity counts-derived PA where SIMEX correction did not show a noticeably stronger association.

The much stronger protective association of LFENMO after age-restricting the external validation data to that of UK Biobank suggests the potential importance of age-related differences in movement patterns captured by accelerometer data. ML did not benefit from the age restriction, potentially because its measurement error structure is independent of participants’ age. However, the results do not necessarily indicate that ML is robust to potential non-generalizability of external validation data to the target cohort without further investigation.

The consistently wider confidence intervals seen in SIMEX-corrected estimates than in the naive estimates are an essential feature of measurement error correction. The larger uncertainty is partly attributable to our novel two-stage bootstrap variance estimator, as observed in the simulation. By accounting for the sampling variance of the validation data, our approach quantifies uncertainty in the dose-response association, thereby preventing potential type I error due to a falsely narrow CI. We note that the smaller (age-subset) validation sample led to substantially wider CIs for associations, an important consideration when collecting ground-truth data for measurement error correction.

Surprisingly, associations estimated from activity counts did not show noticeable changes after SIMEX correction, despite extensive measurement error in this measure. It is possible that selecting other intensity cut-points, or simultaneously integrating multiple cutpoints to the GLS combiner, would lead to entirely different error-correction results. This is because (often arbitrary) choice of cut-points is the primary determinant of accuracy in these methods^11^, hence leading to a recent call to replace cut-point methods with ML.

Extant studies of measurement error largely focused on self-reported PA, demonstrating attenuated etiologic associations^7^. In contrast, accelerometer data were often erroneously assumed to be error-free in etiologic studies^7^, with much of the research focusing on inconsistent prevalence estimates of PA (as opposed to etiologic estimates) due to the selection of intensity cut-points and accelerometer processing methods. The only existing measurement study investigating accelerometer data found that the association between a randomized PA intervention program and the derived PA outcome was attenuated, with ground truth ascertained by doubly labelled water and calorimetry from 106 validation participants^10^. Our work extends this by incorporating non-linear survival analysis, a widely used method in PA research for guideline development^41^.

The use of a pooled PA variable via the GLS combiner may have provided a limited advantage in our study, since the much smaller measurement error of ML dominated the dose-response curve; i.e., the use of ML-derived exposure alone is sufficient. However, we believe that our multivariate method will be useful to perform ensemble error correction by leveraging rapidly increasing numbers of algorithms, device products, and sensor channels (heart rate variability, accelerometer, and Global Positioning System data).

As previously reported, the protective association of PA was noticeably stronger among the female population^3,11^. SIMEX correction further increased this sex-based heterogeneity, as bias correction away from the null was observed only in the female subsample. The result suggests that the current evidence on female-specific benefits of PA may be underestimated. The TPA exposure weighted by Moderate-Vigorous PA showed substantial bias correction (stronger association) even for activity counts, suggesting the potential importance of addressing measurement error in Moderate-Vigorous PA in future investigation.

Our simulation and empirical results indicate that without measurement-error correction, dose-response associations estimated from wearable-derived LPA and TPA may be attenuated, an important consideration for future etiologic studies and the dose-response meta-analyses of existing results. Naive estimates also tend to show narrower CIs when measurement error is not accounted for, which can lead to erroneous conclusions about the amount of LPA and TPA that provide a substantial reduction in the incidence of CVD. Given the increasing availability of validation data for 24-hour movement, the measurement correction should be utilized for dose-response survival analysis.

One of our strengths is using AFT over Cox regression, which increases the transparency of communicating the benefits of PA in terms of time ratios, rather than the proportional hazards model, whose interpretation is challenging for practitioners and the public. While we applied this method to dose-response survival analysis, which is a critical analytical method in PA epidemiology, our AFT-Spline-SIMEX package is applicable to other fields of epidemiology. For instance, it can incorporate multiple instruments on dietary or nutrient intake to model the incidence of nutrition-related chronic diseases through ensemble error correction. As a limitation, our AFT-spline-SIMEX does not currently support competing risk analysis. Survival analyses that account for competing risks and measurement error^42,43^ was demonstrated recently, and we aim to extend this method to accommodate our non-linear analysis. In addition, UK Biobank is a participant-based sample and thus subject to selection bias, even though a previous bias adjustment did not lead to noticeably different associations between PA and CVD.

In addition to the incorporation of competing risks analysis, future work should characterize the measurement error process for Moderate-to-vigorous PA, which may include extensive zero minutes due to the short duration of data collection, e.g., 24 hours or less due to participation burden for wearable camera, though generally non-zero in standard data collection (e.g., 7 days) in cohort.

## Data Availability

Data used in this study are available from UK Biobank to approved researchers upon application. Authors cannot share the data directly.

## Data Availability

UK Biobank data are available to approved researchers through UK Biobank access procedures. Analysis code, R package for Aft-Spline-SIMEX and related documentations are available at [repository/blinded] upon publication.

## Acknowledgments

We appreciate technological support and data preparation by Chi Zhang and Yacine Lapointe.

This research used the NeuroHub infrastructure and was undertaken thanks in part to funding from the Canada First Research Excellence Fund, awarded through the Healthy Brains, Healthy Lives initiative at McGill University.

This research was enabled in part by support provided by Calcul Québec and Compute Canada.

This research has been conducted using the UK Biobank Resource under Application Number 45551.

## Supplementary Material 1: Supplementary Figures and Tables

**Figure S1.**
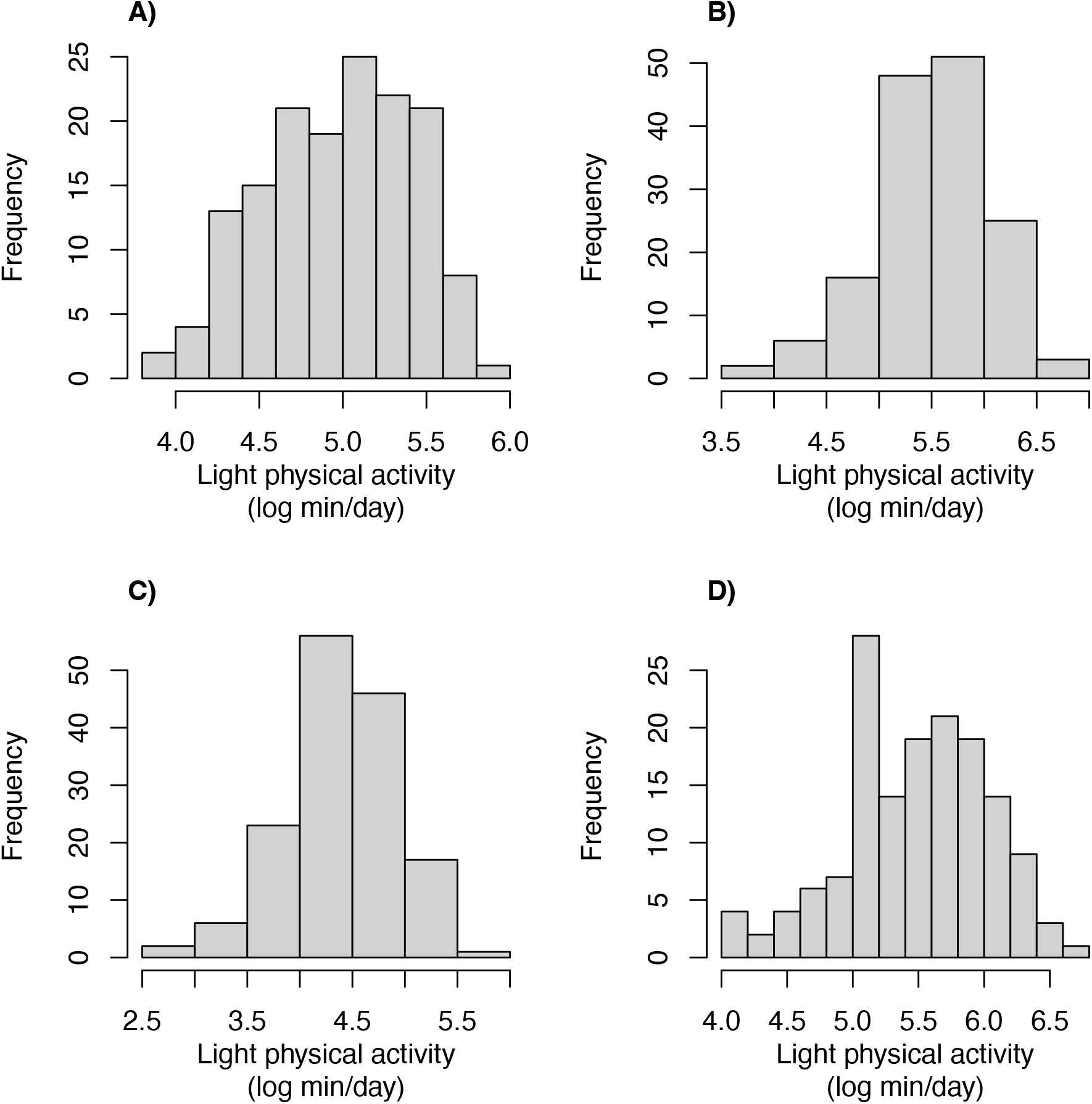
Distribution of log-transformed daily duration of light physical activity measured by **(A)** low-pass filter Euclidean Norm Minus One, **(B)** machine learning, **(C)** activity counts, and **(D)** ground truth.

**Figure S2.**
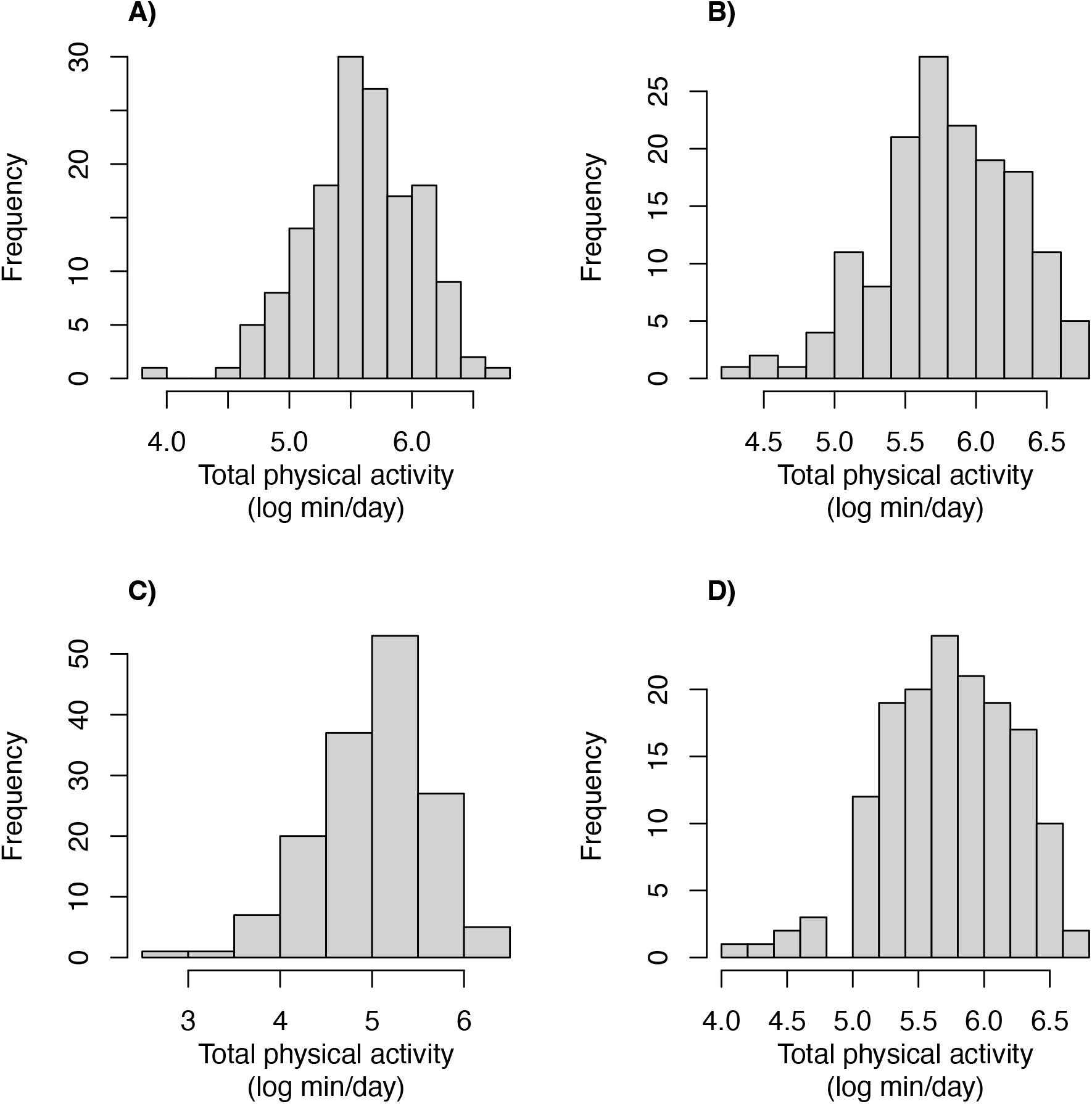
Distribution of log-transformed daily duration of total physical activity measured by **(A)** low-pass filter Euclidean Norm Minus One, **(B)** machine learning, **(C)** activity counts, and **(D)** ground truth.

**Figure S3.**
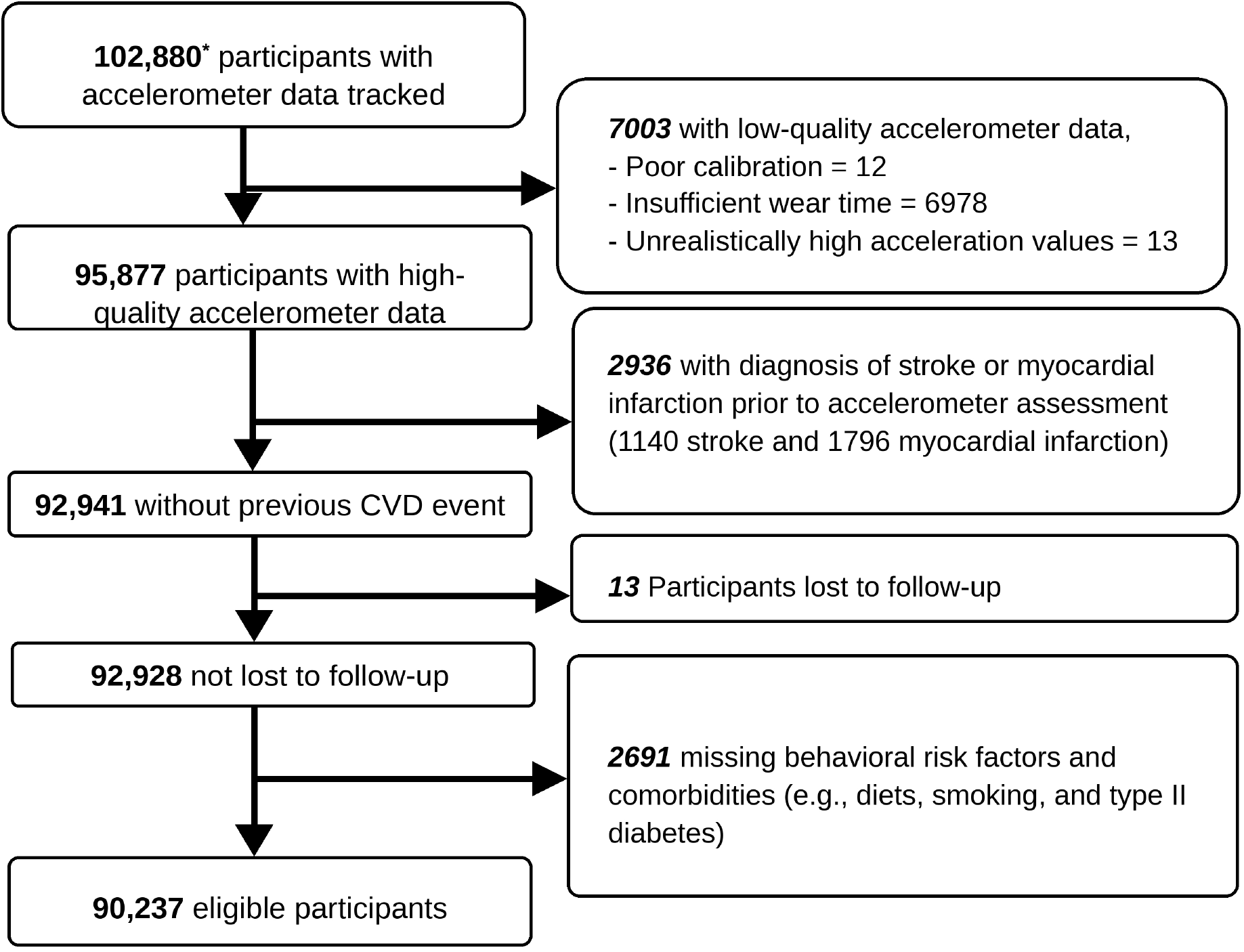
Flowchart describing the exclusion process for adults in the UK Biobank accelerometer sub-cohort, 2013–2022. ∗ This is the remaining number after the UK Biobank removed approximately 1200 individuals who requested to remove their entire data from the cohort. *Abbreviation:* CVD, cardiovascular disease.

**Figure S4.**
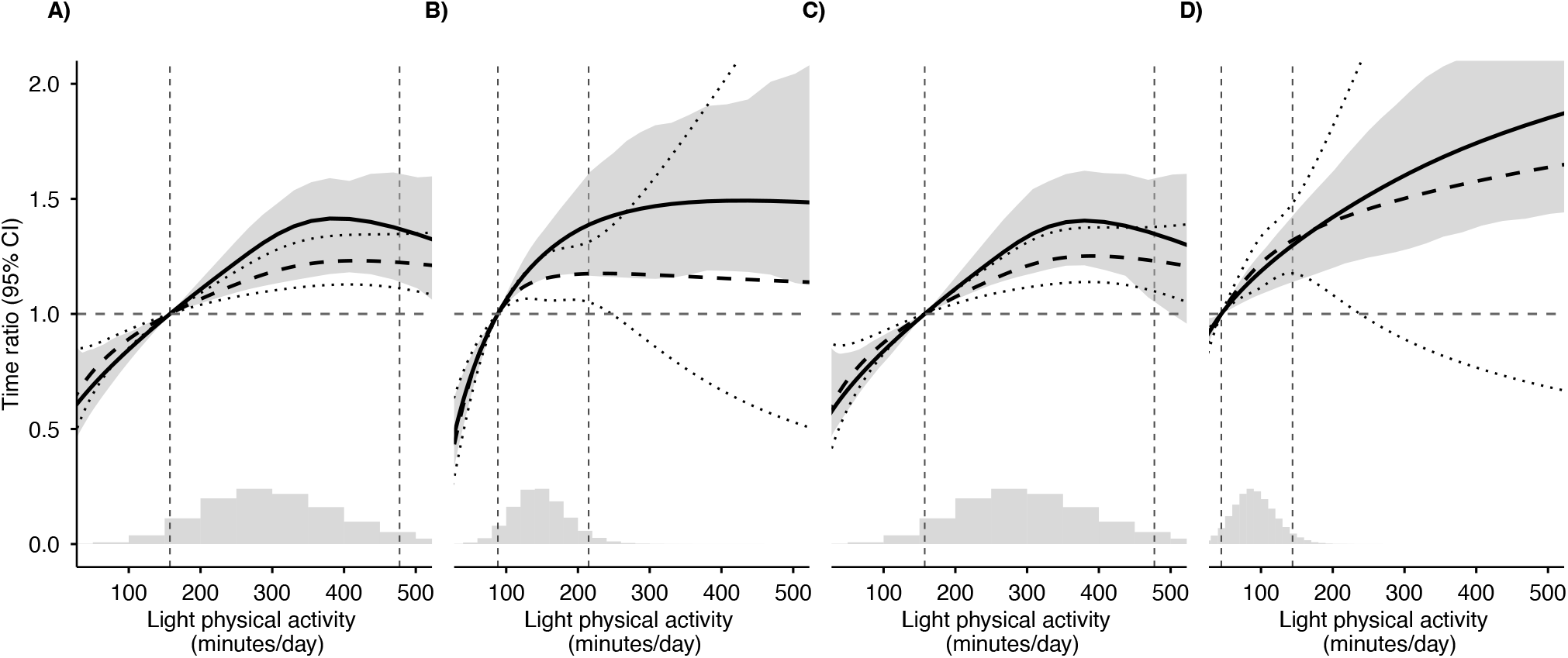
Results based on spline degrees of freedom = 3 for light physical activity. Panels indicate **(A)** Generalized Least Square combined estimate, **(B)** low-pass filter Euclidean Norm Minus One, **(C)** machine learning, and **(D)** activity counts.

**Figure S5.**
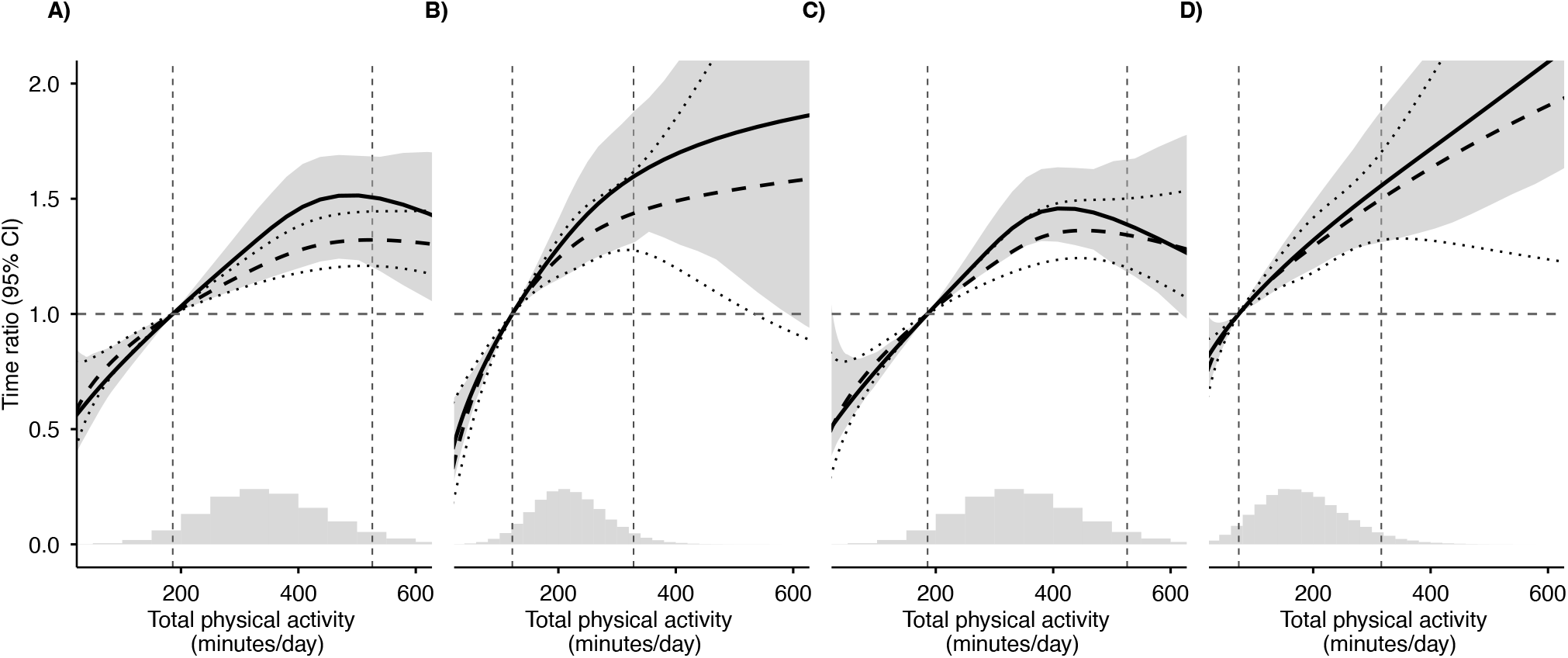
Results based on spline degrees of freedom = 3 for total physical activity. Panels indicate **(A)** Generalized Least Square combined estimate, **(B)** low-pass filter Euclidean Norm Minus One, **(C)** machine learning, and **(D)** activity counts.

**Figure S6.**
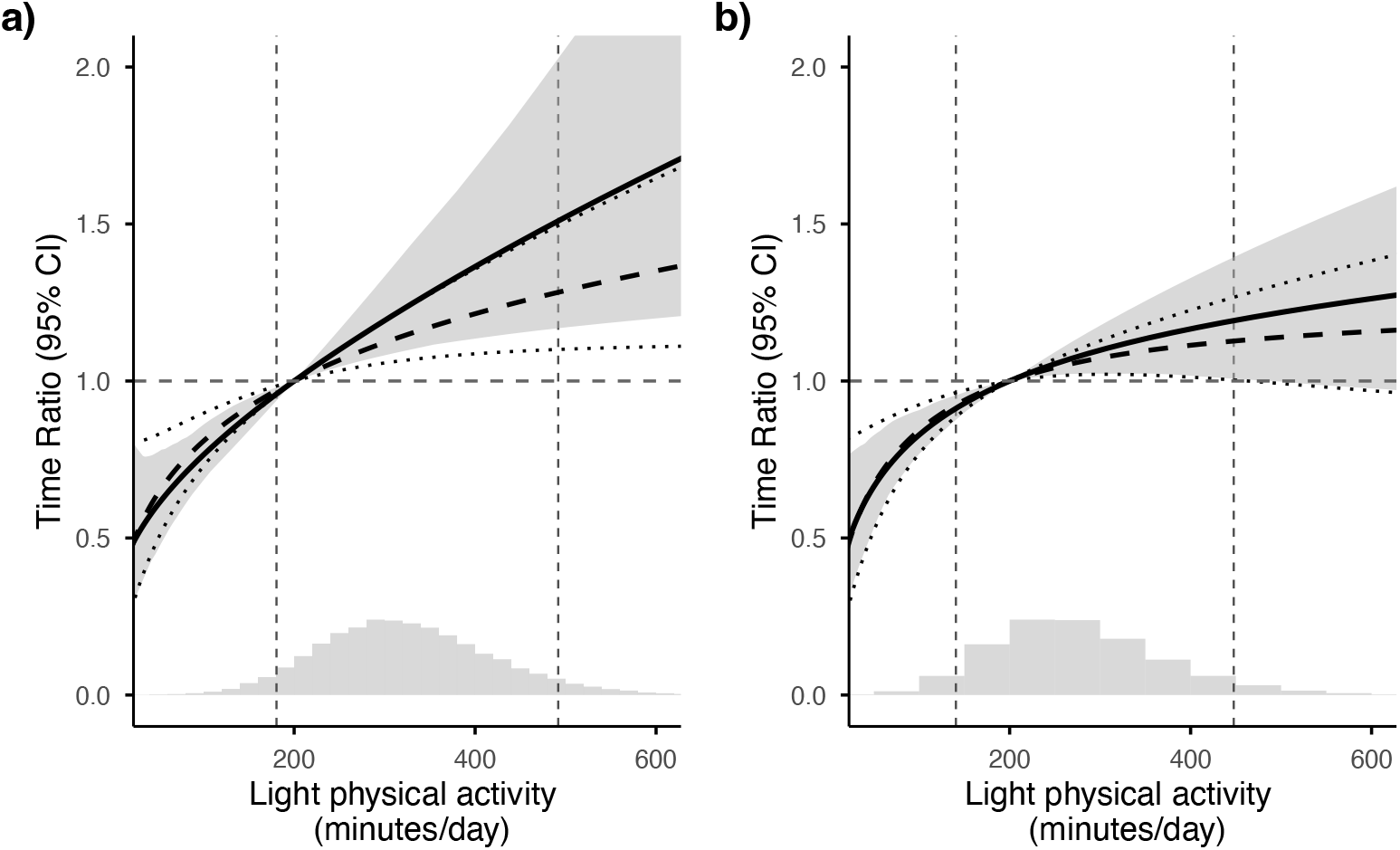
Stratified association of light physical activity by **(A)** female and **(B)** male subsample across SIMEX-corrected (solid line, grey 95% band) and naive (dashed line, dotted 95% CI) estimates.

**Figure S7.**
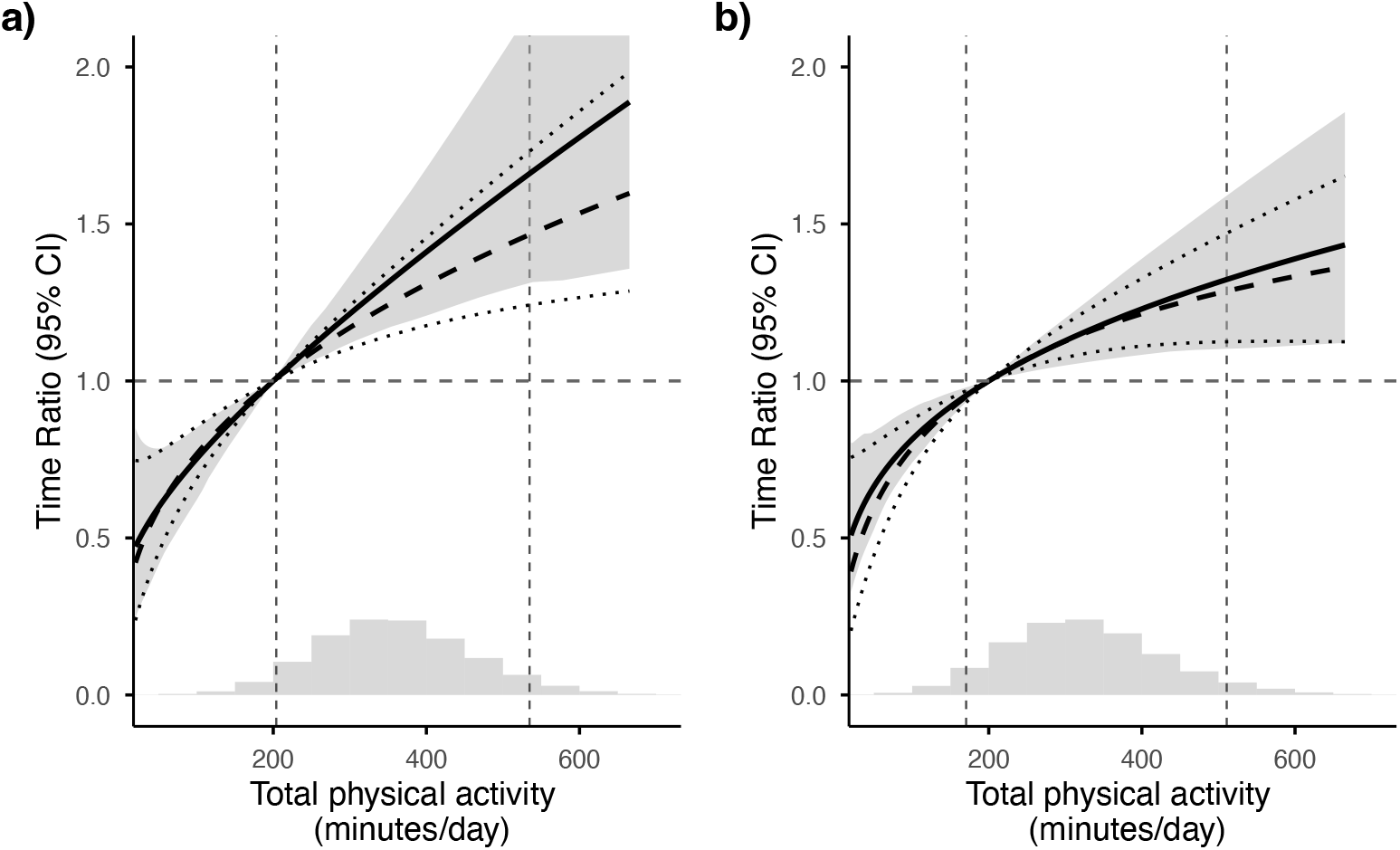
Stratified association of total physical activity by **(A)** female and **(B)** male subsample across SIMEX-corrected (solid line, grey 95% band) and naive (dashed line, dotted 95% CI) estimates.

**Figure S8.**
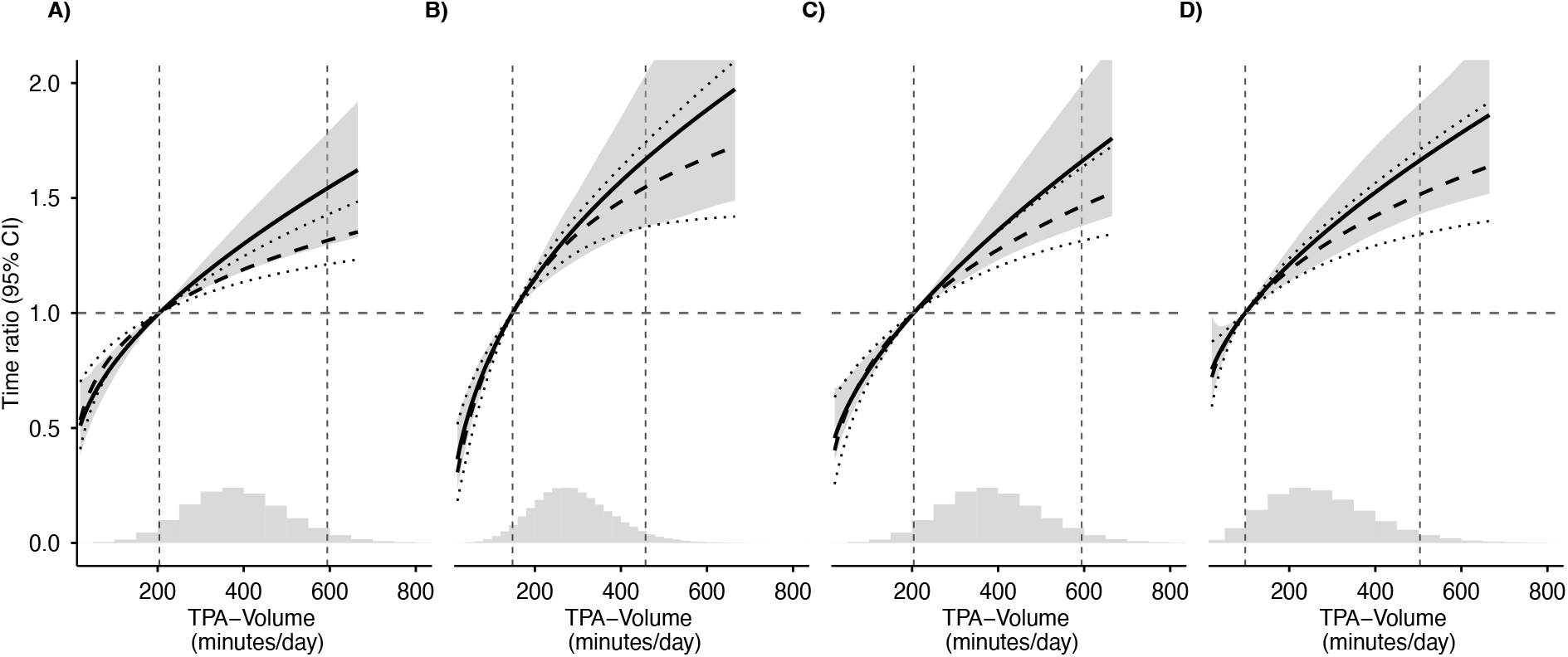
Comparison of dose-response associations for total physical activity weighted by moderate–vigorous physical activity for **(A)** Generalized Least Square combined exposure, **(B)** low-pass filter Euclidean Norm Minus One, **(C)** machine learning, and **(D)** activity counts. SIMEX-corrected curves are solid black with a grey 95% band; naive estimates are dashed with dotted 95% CI.

**Figure S9.**
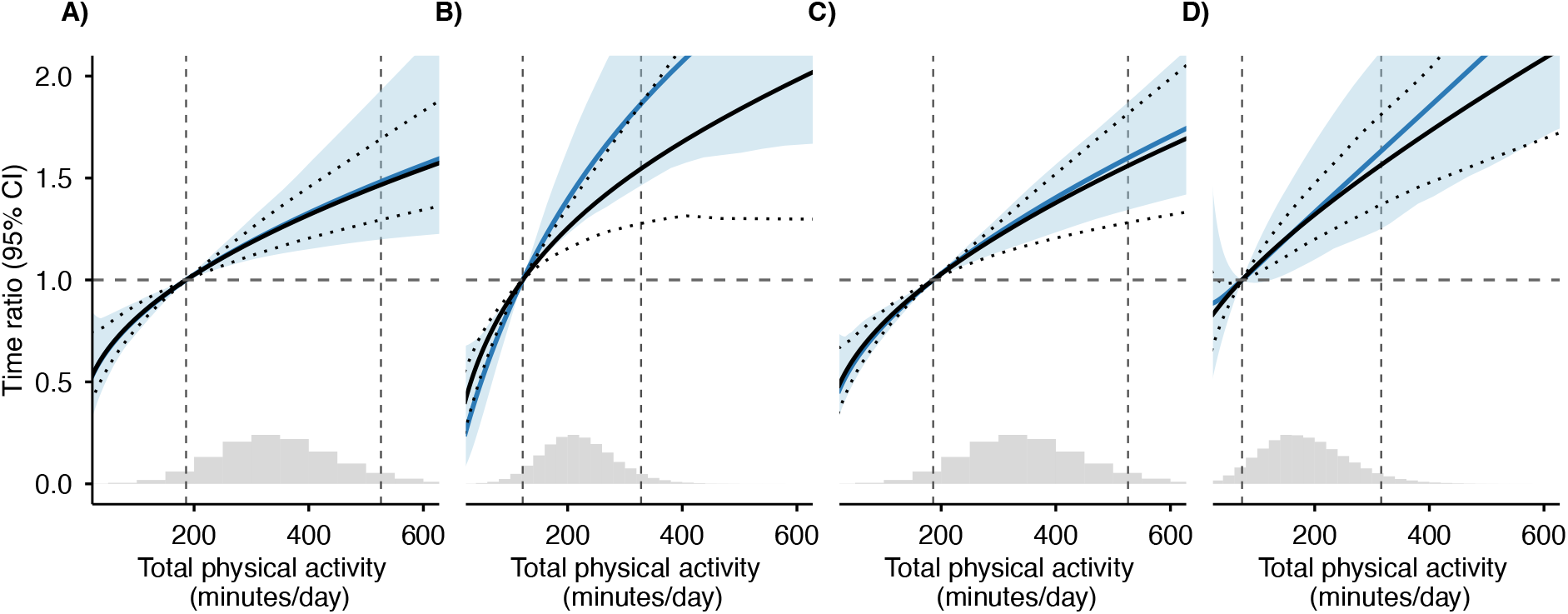
Result of age-restricted validation data to UK Biobank population for total physical activity estimated by **(A)** Generalized Least Square combined exposure, **(B)** low-pass filter Euclidean Norm Minus One, **(C)** machine learning, and **(D)** activity counts. Result from age-restricted validation data is solid blue line light blue shade as 95%CI. Results from the full validation data (main analysis) indicated by black line with dotted lines as 95% CI

**Figure S10.**
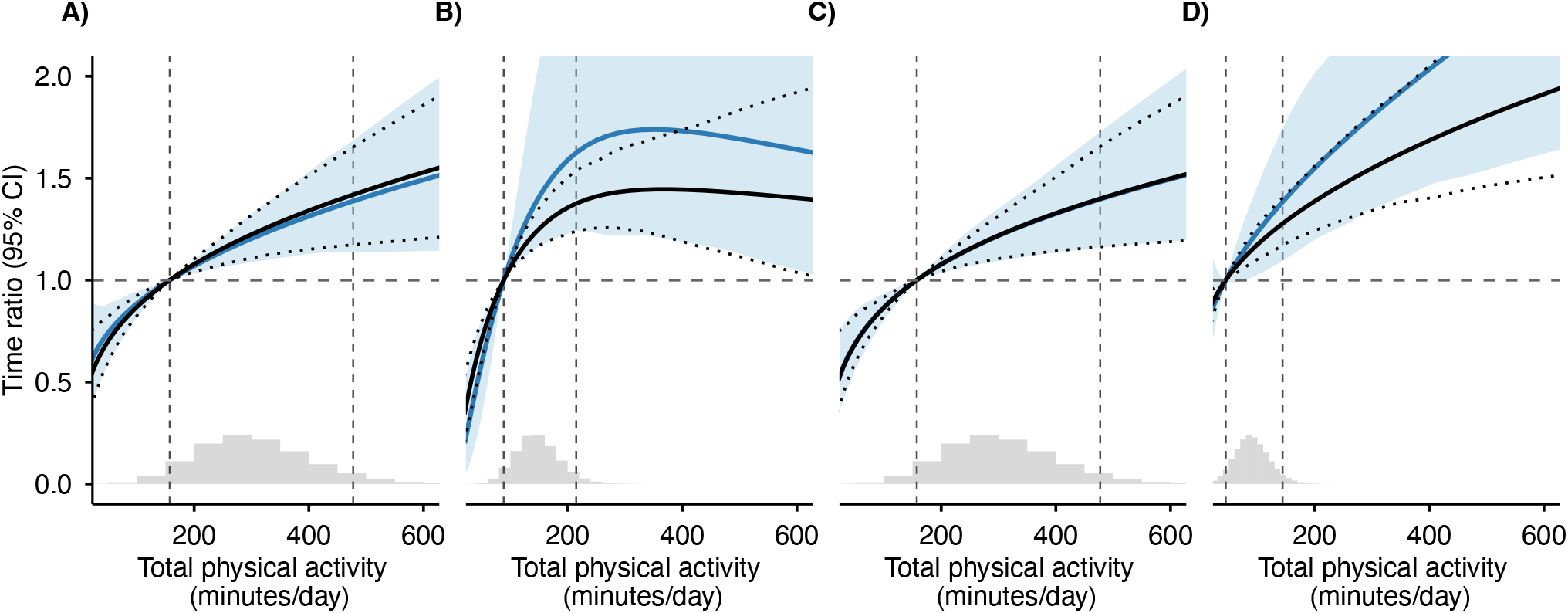
Result of age-restricted validation data to UK Biobank population for light physical activity estimated by **(A)** Generalized Least Square combined exposure, **(B)** low-pass filter Euclidean Norm Minus One, **(C)** machine learning, and **(D)** activity counts. Result from age-restricted validation data is solid blue line light blue shade as 95%CI. Results from the full validation data (main analysis) indicated by black line with dotted lines as 95% CI

## Supplementary Material 2: Details of Statistical Model and Simulation

### Supplementary Appendix S2.1 Accelerated Failure Time Model

Let *T*_*i*_ denote the (potentially right-censored) survival time and *C*_*i*_ the censoring time, so that the observed outcome is (*Y*_*i*_, *δ*_*i*_) = (min(*T*_*i*_, *C*_*i*_), **1**[*T*_*i*_ ≤ *C*_*i*_]). We assume *T*_*i*_ ⊥ *C*_*i*_ | (*W*_*i*_, **Z**_*i*_).

The AFT model is

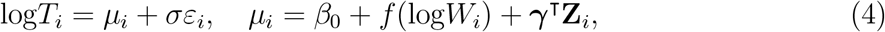

where *σ >* 0 is a scale parameter, *ε*_*i*_ is a standardized error term, and ***θ*** = (*β*_0_, ***β***_*f*_, ***γ***, *σ*) collects all model parameters. The log-likelihood for the observed data 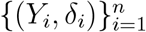 is

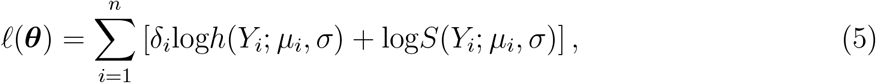

where *h*(·) and *S*(·) are the hazard and survival functions, respectively. Their explicit forms depend on the distributional assumption for *ε*_*i*_.

#### Log-normal AFT

When 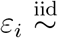 Normal(0, 1), *T* | (*W*, **Z**) follows a log-normal distribution. Letting *φ*(·) and Φ(·) denote the standard normal density and distribution function,

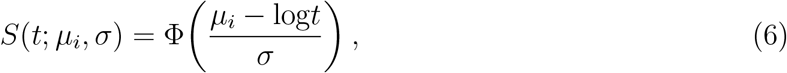

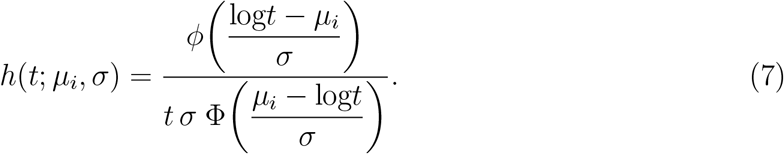

#### Weibull AFT

When *ε*_*i*_ follows the standard extreme-value (Gumbel) distribution, *T*_*i*_ | (*W*_*i*_, **Z**_*i*_) follows a Weibull distribution with scale exp(*μ*_*i*_) and shape 1*/σ*:

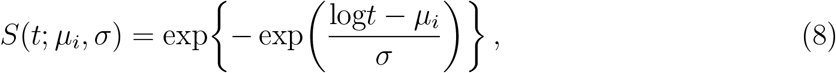

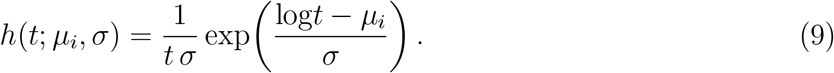

### Supplementary Appendix S2.2 Natural Cubic Spline Basis

The function *f* (·) in (4) is represented as a linear combination of natural cubic spline basis functions. With *K* = 2 interior knots, the spline has *K* + 1 = 3 non-intercept degrees of freedom, so ***β***_*f*_ = (*β*_1_, *β*_2_, *β*_3_)^⊤^ and 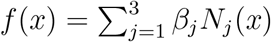.

The interior knots were placed at the 33rd percentile (*ξ*_1_) and the 67th percentile (*ξ*_2_) of the observed 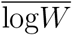 distribution, the default placement of `splines::ns()` at three degrees of freedom. This placement puts approximately equal numbers of observations in each interval between knots, providing locally balanced estimation across the exposure range^44^. The boundary knots *τ*_*L*_ and *τ*_*R*_ were taken as the minimum and maximum of the observed 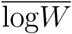 values.

Let 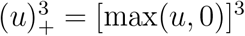 denote the truncated cubic power function. Define

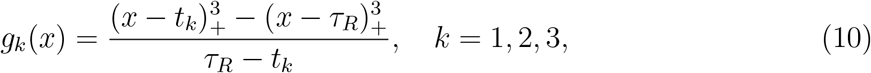

where *t*_1_ = *τ*_*L*_, *t*_2_ = *ξ*_1_, *t*_3_ = *ξ*_2_. The natural cubic spline basis functions are then^44^:

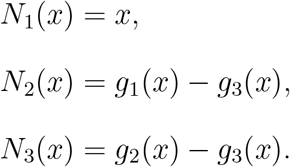

These basis functions satisfy the naturalness constraints: *f* (*x*) is linear for *x< τ*_*L*_ and *x> τ*_*R*_ (i.e., the second derivative vanishes at the boundary knots), and *f* is a cubic polynomial on each interval [*τ*_*L*_, *ξ*_1_], [*ξ*_1_, *ξ*_2_], and [*ξ*_2_, *τ*_*R*_], with continuous first and second derivatives at the interior knots.

### Supplementary Appendix S2.3 Multivariate Measurement Error Model

In practice, the true LPA *W*_*i*_ is unobserved; only the vector of device-derived surrogates 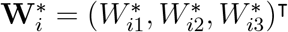 is available. Because the three surrogates are derived from the same raw accelerometer trace, their measurement errors share variation due to wear-time, posture, and signal-processing artefacts. We therefore specify a multivariate log-linear measurement error model,

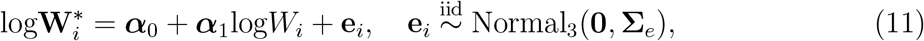

where ***α***_0_, ***α***_1_ ∈ ℝ^3^ are measure-specific intercept and attenuation vectors, **Σ**_*e*_ is a 3 × 3 residual covariance whose off-diagonals capture correlated measurement error, and **e**_*i*_ is independent of *W*_*i*_ and all other covariates. Under model (11), the conditional moments of 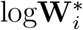 given log*W*_*i*_ are

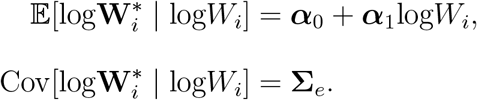

The parameter set (***α***_0_, ***α***_1_, **Σ**_*e*_) was estimated from the Capture-24 validation data by jointly regressing each 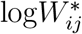 on log*W*_*i*_ via seemingly unrelated regression, with 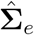 taken as the sample covariance of the residuals. Substituting any single 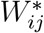 for *W*_*i*_ in the AFT model (4) without correction yields a naive estimator of *f* (·) that is attenuated toward the null due to the presence of *e*_*ij*_; the same attenuation applies to any linear combination of the three surrogates.

### Supplementary Appendix S2.4 GLS Combiner

To pool information across the three surrogates while preserving the classical-error structure required by SIMEX, we combine them using generalized least squares (GLS). First, each surrogate is back-transformed to the scale of log*W*_*i*_ using the Step 1 calibration parameters,

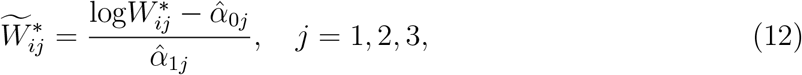

so that 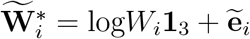, where

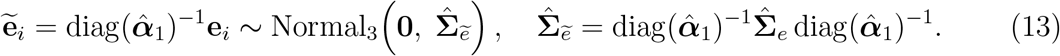

Among all linear combiners 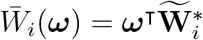 satisfying ***ω***^⊤^**1**_3_ = 1 (so that 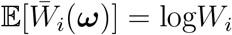), the variance-minimizing choice solves

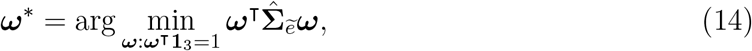

which by Lagrange multipliers gives

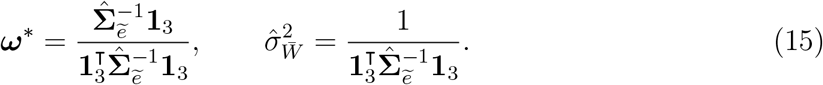

The combined surrogate is 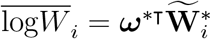, with residual model

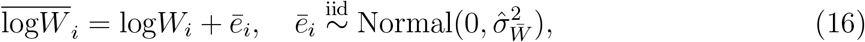

preserving the classical additive error structure on a single combined surrogate. The combiner reduces to the inverse-variance weighting 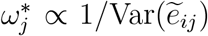 when **Σ**_*e*_ is diagonal, and strictly improves on inverse-variance weighting whenever the off-diagonals are non-zero. Equation (16) is the input to the SIMEX procedure in Appendix Supplementary Appendix S2.5, with 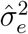 replaced throughout by 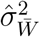.

### Supplementary Appendix S2.5 SIMEX Algorithm

SIMEX corrects for this attenuation by exploiting the known error structure of the combined surrogate (16). The estimator proceeds in two phases: simulation and extrapolation^12,14^.

#### Simulation phase

For each *λ* in a grid Λ and for each replicate *b* = 1,…, *B*, construct a perturbed surrogate

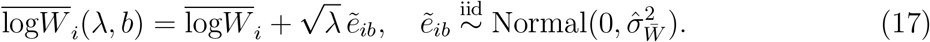

The total measurement error variance in 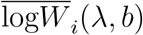 is 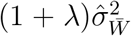, which increases monotonically from 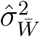 at *λ* = 0 (the observed data) to 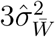 at *λ* = 2. The AFT model (4) is fitted to each perturbed dataset and evaluated on a fixed exposure grid, yielding fitted curves 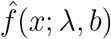. These are averaged across replicates to obtain

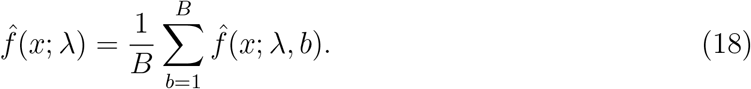

The simulation study used Λ = {0.1, 0.2,…, 2.0} (20 equally spaced values) with *B* = 20; the applied analysis used the coarser Λ = {0.5, 1.0, 1.5, 2.0} with *B* = 100, trading grid resolution for a larger number of perturbed fits at each *λ*.

#### Extrapolation phase

As *λ* increases, the additional measurement error inflates the bias in 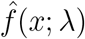 in a smooth and predictable way. At each grid point *x*, the relationship between 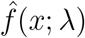 and *λ* is modeled as a quadratic,

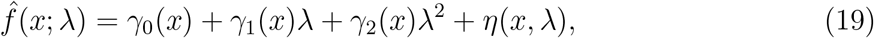

fitted by ordinary least squares across the points *λ* ∈ Λ. The SIMEX-corrected estimate is obtained by evaluating this fitted curve at *λ* = −1, the hypothetical point of zero measurement error,

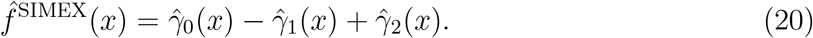

Extrapolating pointwise rather than coefficient-wise matters when the spline is specified by its degrees of freedom, because `splines::ns()` then re-derives the interior knots from whichever dataset it receives. The perturbed exposure in (17) is more dispersed as *λ* grows, so its quantile-based knots move outward with *λ* and the coefficients 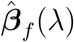 refer to a different basis at each *λ*. The fitted curve 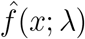 is invariant to that choice of representation, so the pointwise extrapolation is well defined regardless. The two formulations coincide when the knots are held fixed, since *f* is linear in ***β***_*f*_ for a fixed basis. In simulation the two ways of placing knots gave the same accuracy to five decimal places (mean |bias| 0.09482 with knots re-derived at each *λ* against 0.09483 with knots fixed, over 50 replicates), confirming that the correction is insensitive to the drifting basis.

### Supplementary Appendix S2.6 Two-Stage Bootstrap Variance Estimation

The conditional residual variance 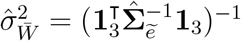 from (15) treats the Step 1 calibration estimates 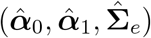 as fixed constants. Be cause these estimates are themselves random functions of the finite validation sample, propagating only 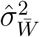 understates the total variability of 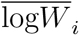, and pointwise confidence intervals for 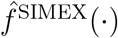 built on this conditional variance alone are anti-conservative. The two-stage nonparametric bootstrap defined below propagates both sources of uncertainty jointly.

Let 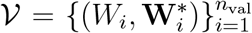 denote the validation dataset and 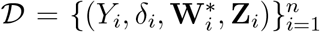 the main analysis dataset. For *r* = 1,…,*R* (with *R* = 200):

1. **Resample the calibration sample**. Draw *V*^(*r*)^ of size *n*_val_ from *V* with replacement.
2. **Refit the multivariate ME model**. Refit (11) on *V*^(*r*)^ to obtain 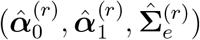. Compute the rescaled covariance 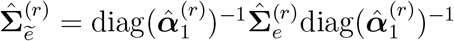, the GLS weights

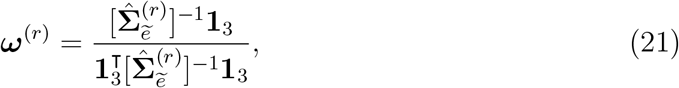

and the bootstrap-replicate residual variance 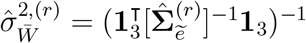.
3. **Resample the main sample**. Draw D^(*r*)^ of size *n* from D with replacement.
4. **Form the bootstrap combined surrogate**. For each participant *i* ∈ D^(*r*)^, compute 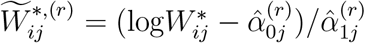 and 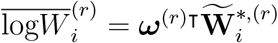.
5. **Apply SIMEX**. Run the full SIMEX procedure (Section Supplementary Appendix S2.5) on D^(*r*)^ using the bootstrap combined surrogate from step 4 and the bootstrap conditional variance 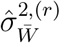 from step 2 in the simulation step (17). This yields 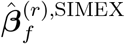 and the corrected dose-response curve 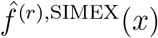.

The pointwise 95% confidence interval at *x* is 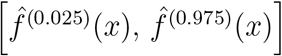, where 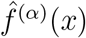 denotes the *α*-quantile of 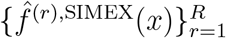. The two stages are resampled independently, consistent with the design of an external validation study in which Capture-24 and UKBB participants are non-overlapping. Under standard regularity conditions, this nested bootstrap is asymptotically equivalent to the sandwich variance of the joint M-estimator stacking Step 1 and Step 2 estimating equations^13^, with the advantage of not requiring closed-form expressions for the influence functions of the SIMEX-corrected dose-response curve.

### Supplementary Appendix S2.7 Simulation study

**Table S1.** Monte Carlo summary of the integrated squared error (ISE) across *R* = 500 replicates of the simulation described in Appendices Supplementary Appendix S2.7.1– Supplementary Appendix S2.7.3. MCSE denotes the Monte Carlo standard error of the mean.

| Estimator | Mean ISE | MCSE | Median | 2.5% | 97.5% |
| --- | --- | --- | --- | --- | --- |
| Oracle | 0.018 | 0.001 | 0.011 | 0.003 | 0.068 |
| Naive | 0.077 | 0.002 | 0.073 | 0.018 | 0.157 |
| SIMEX | 0.031 | 0.001 | 0.022 | 0.004 | 0.104 |

**Figure S11.**
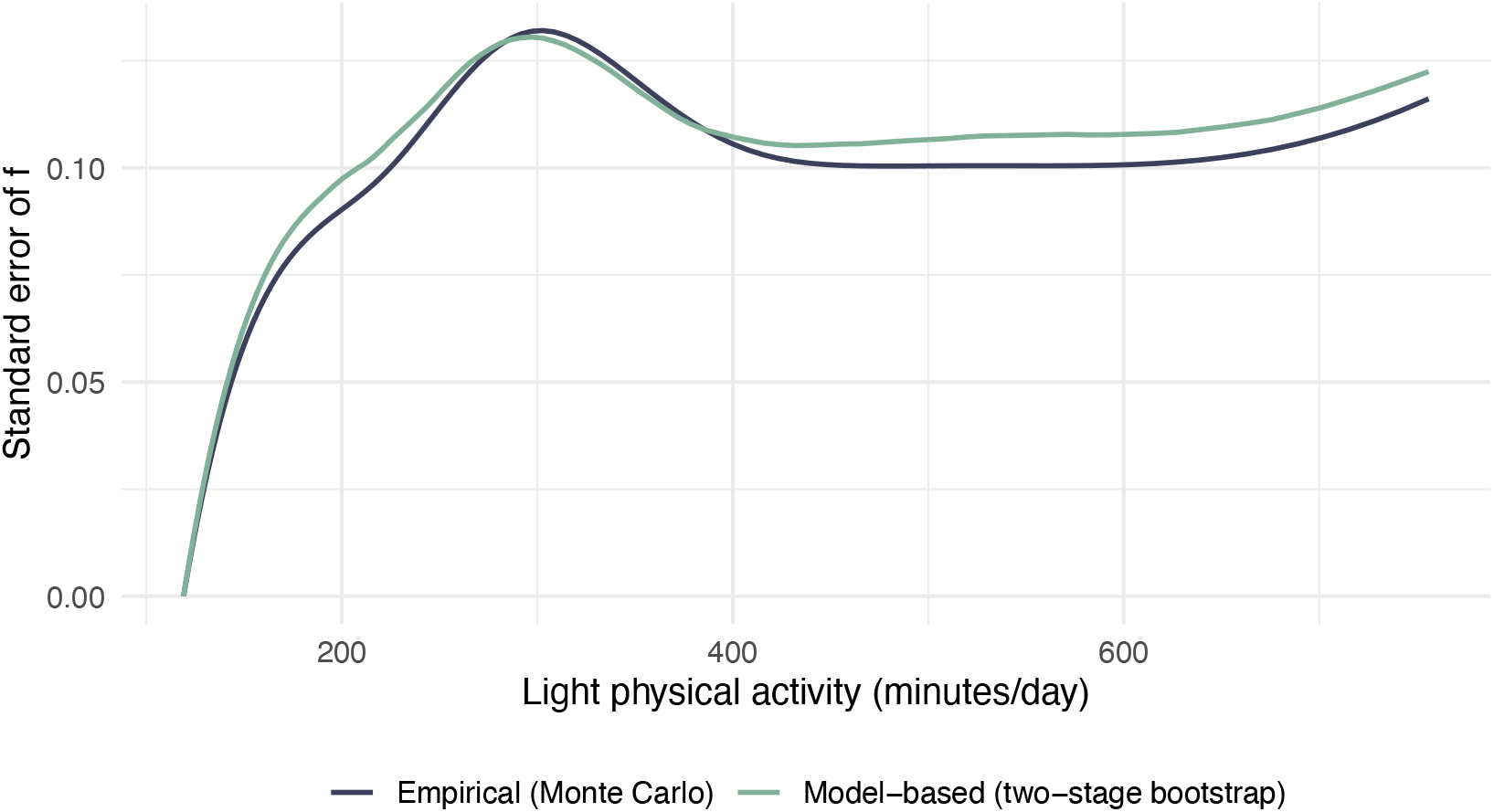
Empirical (Monte Carlo) standard error and model-based (two-stage bootstrap) standard error of the SIMEX-corrected curve as functions of daily LPA in minutes. The two summaries track one another across the grid (mean ratio ≈ 1.05, the bootstrap if anything marginally conservative), indicating that the bootstrap recovers the sampling variability of 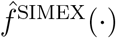 honestly. The pointwise coverage shortfall reported in this subsection therefore reflects point-estimate bias rather than under-stated uncertainty.

#### Supplementary Appendix S2.7.1 Data-generating process

We conducted a Monte Carlo simulation to evaluate the finite-sample performance of the AFT–spline–SIMEX estimator under classical measurement error in a nonlinear exposure.

**Figure S12.**
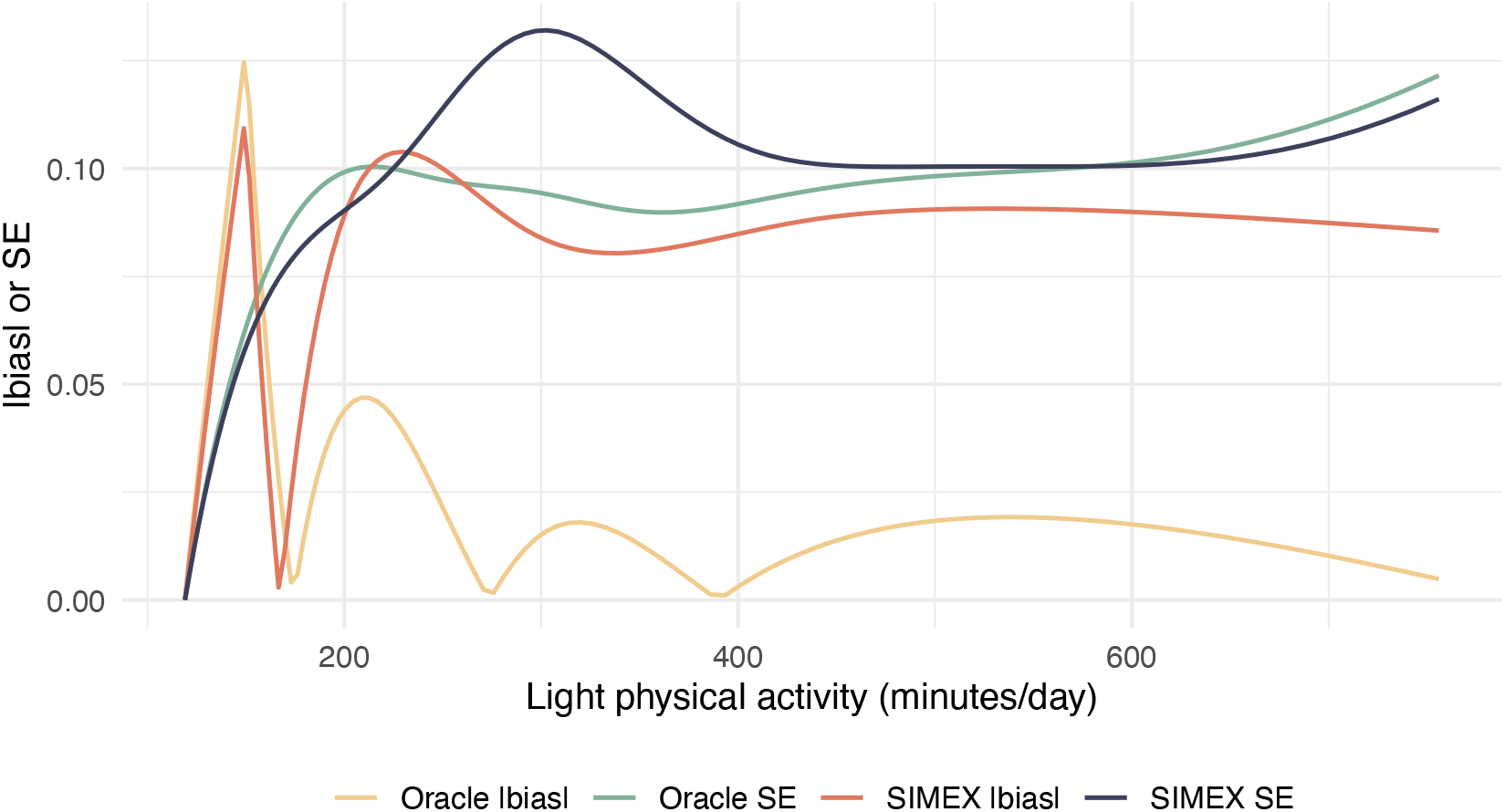
Pointwise absolute bias and empirical standard error of the SIMEX and Oracle estimators against daily LPA in minutes. The SIMEX bias exceeds the Oracle (spline-smoothing) bias across much of the support (the Oracle accounts for only about one-third of it), so at this realistic measurement-error level the residual bias is predominantly SIMEX-specific extrapolation. The SIMEX bias is comparable to its standard error in the steep region, which is the source of the coverage shortfall.

For each replicate, *n* = 2000 participants were generated. Let *W*_*i*_ denote the true exposure on the analysis (log) scale of daily light physical activity. We drew *W*_*i*_ ∼ Normal(log300, 0.20), that is, with mean log300 ≈ 5.70 and variance 0.20 (standard deviation 0.45), so that the latent daily LPA exp(*W*_*i*_) has a median of 300 minutes with 95% of participants between roughly 124 and 725 minutes, a realistic daily range that respects the physiological ceiling, in contrast to an arbitrary-unit scale. The confounder vector **Z**_*i*_ = (*Z*_*i*1_, *Z*_*i*2_, *Z*_*i*3_, *Z*_*i*4_)^⊤^ was drawn with 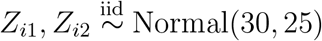, *Z*_*i*3_ ∼ Bernoulli(0.7), and *Z*_*i*4_ ∼ Bernoulli(0.8), all mutually independent.

Event times were generated from a log-normal AFT model,

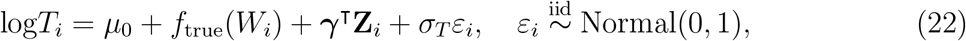

with *μ*_0_ = 4, *σ*_*T*_ = 0.8, and ***γ*** = (0.10, −0.05, 0.30, −0.20)^⊤^. The true dose-response curve was specified as a saturating exponential,

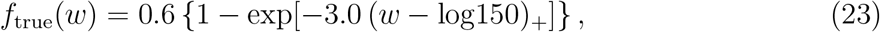

where (*u*)_+_ = max(*u*, 0). This shape encodes a threshold at log150 (150 minutes/day) followed by diminishing returns that saturate by roughly 300 minutes/day, mimicking the saturating association expected between LPA and incident MI. Independent right-censoring times were drawn as *C*_*i*_ ∼ Exp(0.0005), calibrated to yield an event rate of approximately 76% at the chosen *μ*_0_, ***γ***, and *σ*_*T*_ . The observed outcome was (*Y*_*i*_, *δ*_*i*_) = (min(*T*_*i*_, *C*_*i*_), **1**[*T*_*i*_ ≤ *C*_*i*_]).

A vector of three surrogate measures was generated from a classical additive multivariate error model,

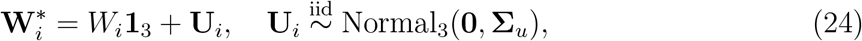

whose marginal error variances were scaled to a target mean per-surrogate reliability of 0.50, where reliability is the intraclass ratio 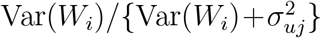, while preserving the relative measurement qualities 2.0 : 2.5 : 3.0 across the three channels, giving 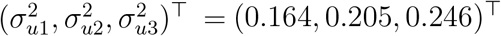, with a common pairwise correlation *ρ* = 0.4 reflecting the shared accelerometer signal of the three measures (Activity Count, Vector Magnitude, ML-derived). Anchoring the error magnitude to a reliability target (rather than to fixed variances) keeps the measurement error commensurate with the exposure variance on the daily scale, where an unscaled choice would render the surrogates nearly pure noise. The target of 0.50 sits at the low end of the reliability range over which the estimator is intended to operate and was chosen deliberately as a stress test: it implies that half the variance of each surrogate is measurement error, a more severe level of contamination than the agreement observed for the best-performing accelerometer processing method in the Capture-24 validation data. The simulation results below should therefore be read as a conservative bound on the behaviour of the estimator in the applied analysis rather than as a match to it. The pre-calibration coefficients were fixed at ***α***_0_ = **0**_3_ and ***α***_1_ = **1**_3_, so each surrogate already lies on the scale of *W*_*i*_ (no Step 1 back-transform needed). An external validation set of *n*_val_ = 500 with paired 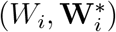 was generated from the same joint distribution to estimate 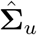 and supply it to the SIMEX procedure.

#### Supplementary Appendix S2.7.2 Simulation study: estimators

We compared three estimators of the dose-response curve *f* (·):

1. **Oracle**. The AFT–spline model (4) fitted on the true exposure *W*_*i*_. This estimator is not feasible in practice and serves as the best-attainable benchmark.
2. **Naive**. The AFT–spline model fitted on the GLS-combined surrogate (Appendix Supplementary Appendix S2.4),

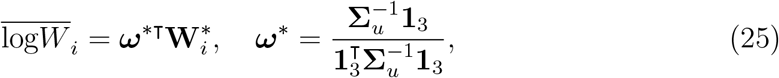

whose residual error variance is 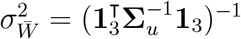. No measurement-error correction is applied.
3. **SIMEX**. The AFT–spline–SIMEX estimator (Appendix Supplementary Appendix S2.5) applied to 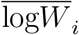 with assumed error variance 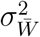, error inflation grid *λ* ∈ {0.1, 0.2,…, 2.0} (20 equally spaced values), *B* = 20 simulation replicates per *λ*, and quadratic extrapolation to *λ* = −1. Pointwise 95% confidence intervals were obtained from *R* = 200 nonparametric bootstrap replicates of the full SIMEX procedure (refitting the GLS combiner on each bootstrap sample).

All spline fits used natural cubic splines with 4 degrees of freedom via `splines::ns()` and were fitted by `survival::survreg()`.

#### Supplementary Appendix S2.7.3 Simulation Study:Performance Metric

Each fitted curve was centered at the lower end of the evaluation grid, 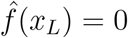, to remove unidentifiability with the AFT intercept. Performance was assessed by the integrated squared error,

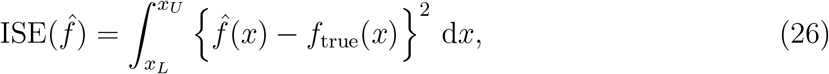

computed by trapezoidal quadrature on a grid of 100 equally spaced values between the 2nd and 98th percentiles of the true exposure distribution. Averages of the ISE across Monte Carlo replicates, together with empirical curve coverage of the bootstrap pointwise 95% intervals, provide the primary summaries of estimator accuracy.

#### Supplementary Appendix S2.7.4 Simulation Study: Results

Across *R* = 500 Monte Carlo replicates, the censoring scheme delivered a mean event rate of 75.8%, and the estimated GLS weights 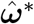 averaged (0.481, 0.311, 0.208)^⊤^, closely tracking the inverse-variance ordering of **Σ**_*u*_, with mean residual variance 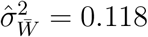 on the log scale. Table S1 summarizes the integrated squared error (ISE) of the three estimators, and Figure 1 displays the corresponding mean curves with pointwise 2.5%–97.5% empirical envelopes.

The naive estimator carries roughly 4.3 times the mean ISE of the oracle (0.077 versus 0.018), confirming that ignoring the additive multivariate measurement error in the surrogate channels induces substantial attenuation of the dose-response curve even after GLS combination. Applying SIMEX restores most of the lost accuracy: the mean ISE drops to 0.031, removing approximately 78% of the naive estimator’s excess error relative to the oracle. The improvement is consistent across the distribution. The SIMEX median ISE (0.022) and upper quantile (0.104) are likewise much closer to the oracle than to the naive baseline and the Monte Carlo standard errors confirm that these differences are not attributable to sampling noise. Figure 1 shows the same pattern visually: the naive mean curve is systematically flatter than the truth across the interior of the exposure support, while the SIMEX mean curve aligns closely with the truth and with the oracle.

#### Supplementary Appendix S2.7.5 Simulation Study: Coverage and Bias Decomposition

The pointwise 95% interval produced by the two-stage bootstrap for the SIMEX-corrected curve attained an empirical coverage of 85.0% across the full evaluation grid and 85.1% in the interior region between the 5% and 95% quantiles of *X*, falling short of the nominal level by approximately ten percentage points. Two mechanisms could in principle account for this shortfall: an under-estimated standard error, or a biased point estimate around which an otherwise correctly sized interval is mis-centered. The two-stage bootstrap explicitly propagates Step 1 and Step 2 sampling variation, but its calibration depends on the bootstrap distribution mimicking the sampling distribution of 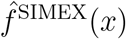, which is not guaranteed for nonlinear plug-in estimators of this form.

To distinguish the two mechanisms, we compared the model-based standard error implied by the bootstrap interval against the empirical standard error of 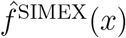 across independent Monte Carlo replicates. At each grid point we computed 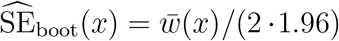, where 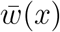 is the mean pointwise interval width across the *R* = 100 outer replicates of the coverage study, and contrasted it with the Monte Carlo standard deviation of 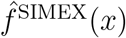 computed across the independent *R* = 500 replicates summarized in Table S1. The two summaries agree closely: average 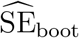 was 0.098 on the full grid and 0.102 in the interior, while the empirical standard error was 0.094 and 0.098 respectively, yielding 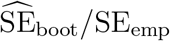 ratios of 1.05 on both. Figure S11 overlays the two curves and shows them to track one another across the support of *X*. The two-stage bootstrap is therefore not anti-conservative in the variance sense (if anything marginally conservative), so the coverage shortfall reflects bias in the point estimate rather than under-stated uncertainty.

We next quantified the bias contribution. The pointwise absolute bias of the SIMEX curve averages 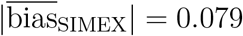 across the grid, comparable in magnitude to its standard error (bias-to-SE ratio 0.84), which is what mis-centers the otherwise correctly sized interval. The Oracle estimator, which by construction conditions on the true exposure *X* and is exempt from measurement-error correction, exhibits a smaller average 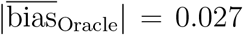 (bias-to-SE ratio 0.30) arising purely from natural-cubic-spline smoothing of the steep rising segment of *f*_true_. Approximately one-third (34%) of the SIMEX bias is therefore shared with the Oracle as spline-smoothing bias; the remaining ∼ 0.05 (about two-thirds) is SIMEX-specific extrapolation bias. This is the expected behaviour at a deliberately conservative measurement-error level: at reliability 0.50 the attenuation that SIMEX must undo is large, so the quadratic extrapolation back to *λ* = −1 travels further and leaves a larger residual bias than under the milder error of the previous arbitrary-unit scenario, in which the Oracle accounted for about two-thirds (rather than one-third) of the total bias. The SIMEX-specific component therefore shrinks as reliability improves, so the coverage reported in this subsection is a floor rather than the level expected at the agreement observed in Capture-24. Figure S12 contrasts the pointwise absolute bias and standard error budgets for the two estimators across the support of *X*.

A sensitivity analysis varying the spline degrees of freedom over df ∈ {2, 3, 4, 5, 6, 8} on the Oracle fit (*N* = 50 replicates) shows that the smoothing-bias component is modest across the whole range and trades off against variance in the usual way. Average |bias| is essentially flat at 0.031 for df = 2, 3 and 4, reaches a minimum of 0.016 at df = 6, and rises to 0.022 at df = 8, while the pointwise standard deviation increases monotonically with complexity, from 0.078 at df = 2 to 0.109 at df = 8. Thus df = 6 minimizes bias and df = 2 minimizes variance. Importantly, the specification used in the applied analysis, df ∈ {2, 3}, carries the same average smoothing bias as the df = 4 fit used for the simulation summaries; its penalty is a larger local bias at the steepest part of the curve (maximum |bias| of 0.153 versus 0.113 at df = 4). Across the whole range the smoothing bias remains small relative to the SIMEX-specific component quantified above. We retain df = 4 as the prespecified choice for the simulation summaries for comparability. The applied analysis uses df = 2 (one interior knot) as its primary specification and reports df = 3 (two interior knots) as a sensitivity analysis; the two fits were not distinguishable by AIC (ΔAIC *<* 4). The one-knot model was chosen in advance on subject-matter grounds, that dose-response curves for physical activity are typically smooth and monotone with diminishing returns rather than richly featured, and the df sweep above is consistent with that choice: over df ∈ {2, 3, 4} the mean absolute bias is flat while the pointwise variance is smallest at df = 2, so the additional flexibility buys no accuracy on a curve of this shape and costs precision. Figure S13 displays the pointwise Oracle bias across df levels.

**Figure S13.**
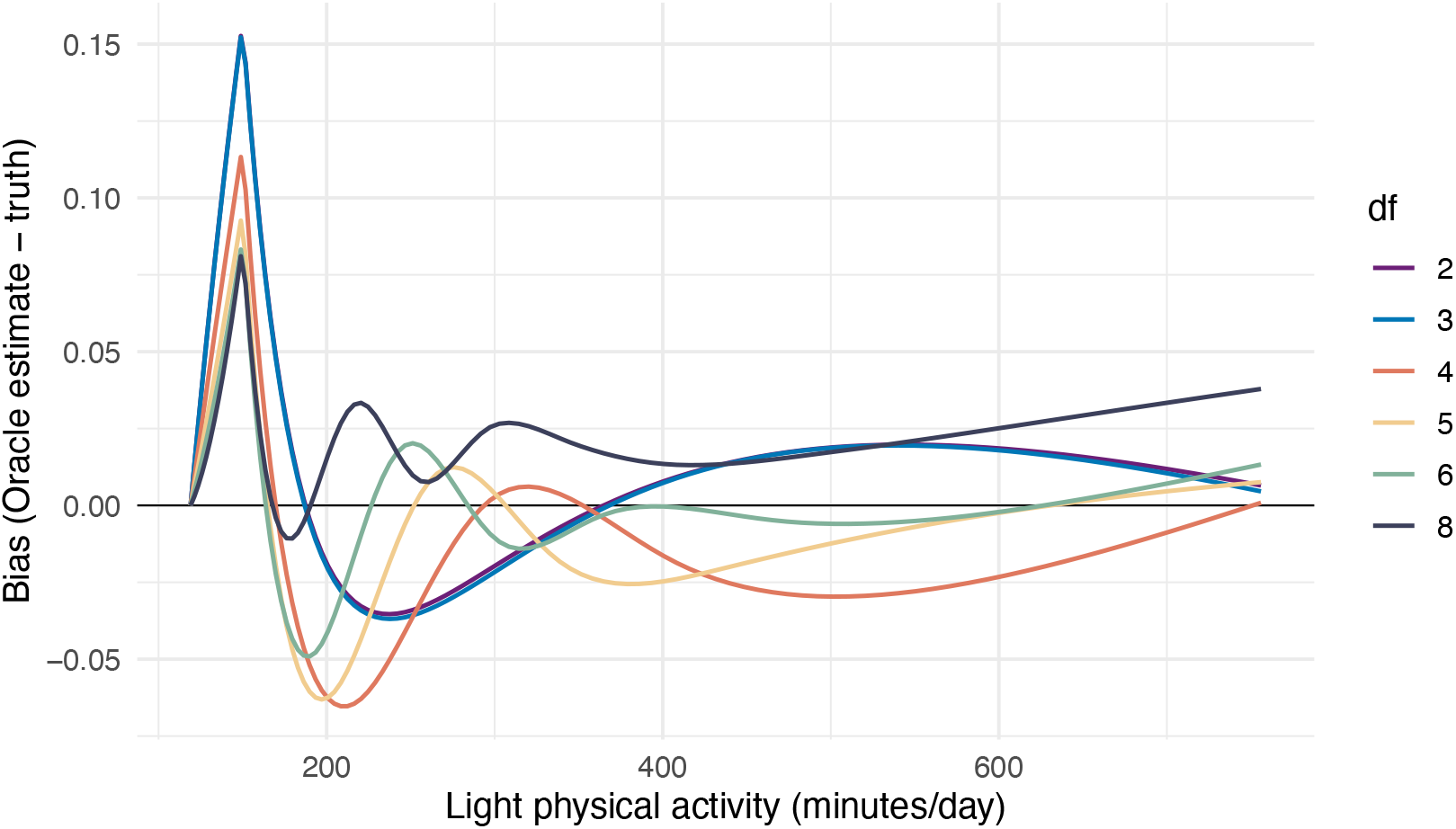
Pointwise bias of the Oracle estimator against daily LPA in minutes across spline degrees of freedom df ∈ {2, 3, 4, 5, 6, 8} (*N* = 50 replicates). Degrees of freedom 2 and 3 are the specifications used in the applied analysis. The smoothing-bias component is small relative to the SIMEX-specific extrapolation bias quantified in this subsection at every level.

The overall reading of this decomposition is that SIMEX delivers a large accuracy gain whose uncertainty is honestly quantified, at the cost of a residual point-estimate bias. On the one hand, SIMEX removes 78% of the naive attenuation on the ISE scale (Table S1) and the two-stage bootstrap recovers its sampling variability faithfully (SE ratio ≈ 1.05). On the other hand, at this deliberately conservative measurement-error level the residual SIMEX bias is majority extrapolation-driven rather than spline-driven, and it is of the same order as the standard error, which leaves the pointwise interval mildly anti-conservative (∼ 85% coverage). We therefore present the SIMEX curve as a substantially de-attenuated point estimate, much closer to the truth than the naive curve, whose pointwise intervals are honest in width but should be read as mildly anti-conservative near the steep region, and we emphasize interpretation over the well-supported interior of the exposure (Appendix Supplementary Appendix S2.7.7).

#### Supplementary Appendix S2.7.6 Simulation Study: Value of the Validation-Aware Two-Stage Interval

A central methodological feature of our approach is that the two-stage bootstrap resamples the *external validation sample* in addition to the main study, thereby propagating the Step 1 calibration uncertainty that a conventional one-stage interval, which holds the calibration fixed, ignores. To quantify the value of this validation-aware design, we reran the coverage study comparing the two-stage interval against the one-stage (conventiona, naive boostrap) interval across a range of validation-sample sizes *n*_val_ ∈ {500, 150, 50} at fixed *n* = 2000 (*N* = 50 outer replicates, *R* = 120 inner bootstraps). The contrast is governed by the size of the validation sample: Step 1 uncertainty is *O*(1*/n*_val_), so it contributes a share of the total variance that grows as the validation sample shrinks.

Table 1 shows that when the validation sample is large relative to the main study (*n*_val_ = 500, ratio 4) the two designs are practically indistinguishable, but as *n*_val_ shrinks the one-stage interval increasingly under-covers and narrows: at *n*_val_ = 50 (ratio 40) one-stage coverage falls to 0.83 while the two-stage holds 0.90, and the two-stage interval is 1.25 times wider. Both designs remain below the nominal level, because both inherit the same biased SIMEX point estimate quantified in Appendix Supplementary Appendix S2.7.5. The informative comparison is therefore the difference between them, which widens monotonically as the validation sample shrinks, from 1.6 to 2.4 to 7.2 percentage points. The effect is clearest at a deliberately small validation sample: at *n*_val_ = 25 (with *n* = 15,000), the two-stage interval was 2.2 times wider than the one-stage interval over the central 90% of the exposure (Figure 2 in the main text). Capture-24 is considerably larger at *n*_val_ = 151, close to the *n*_val_ = 150 row of Table 1, where the width inflation is 1.07. How much of the penalty is realised in any particular dataset depends not on the sample sizes alone but on how precisely the calibration itself is determined, which is a property of the validation study. The value of the validation-aware design is that it prices that uncertainty rather than assuming it away.

#### Supplementary Appendix S2.7.7 Simulation Study: Boundary Behavior of the SIMEX Curve

A characteristic feature of the SIMEX-corrected curve is mild instability near the upper end of the exposure grid, where the SIMEX estimate may droop slightly below the truth even as the naive curve continues to plateau. SIMEX deconvolves attenuation by fitting the model at *λ* ∈ {0.1, 0.2,…, 2.0} and extrapolating quadratically to *λ* = −1. In regions where the surrogate distribution is sparse, the perturbed-data fits become high-variance, and the quadratic extrapolation amplifies that variance. Interpretation of the SIMEX curve should accordingly emphasize the interior region of the exposure support, and the pointwise bootstrap intervals, which widen markedly at the boundaries, provide a transparent visual signal of where the correction is well-supported by the data.

## Supplementary Material 3. STROBE Checklist

STROBE Statement—checklist of items that should be included in reports of observational studies.

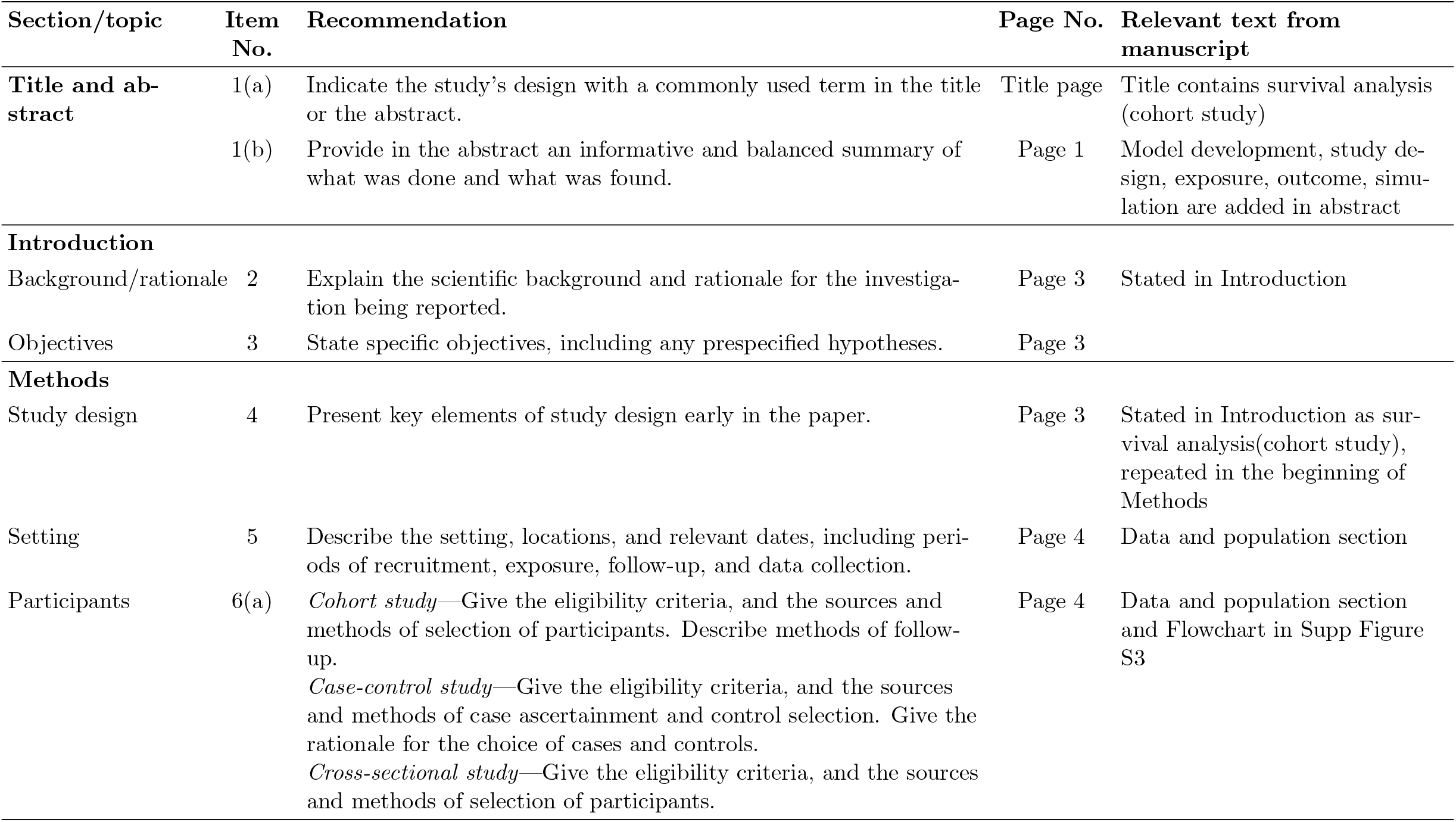
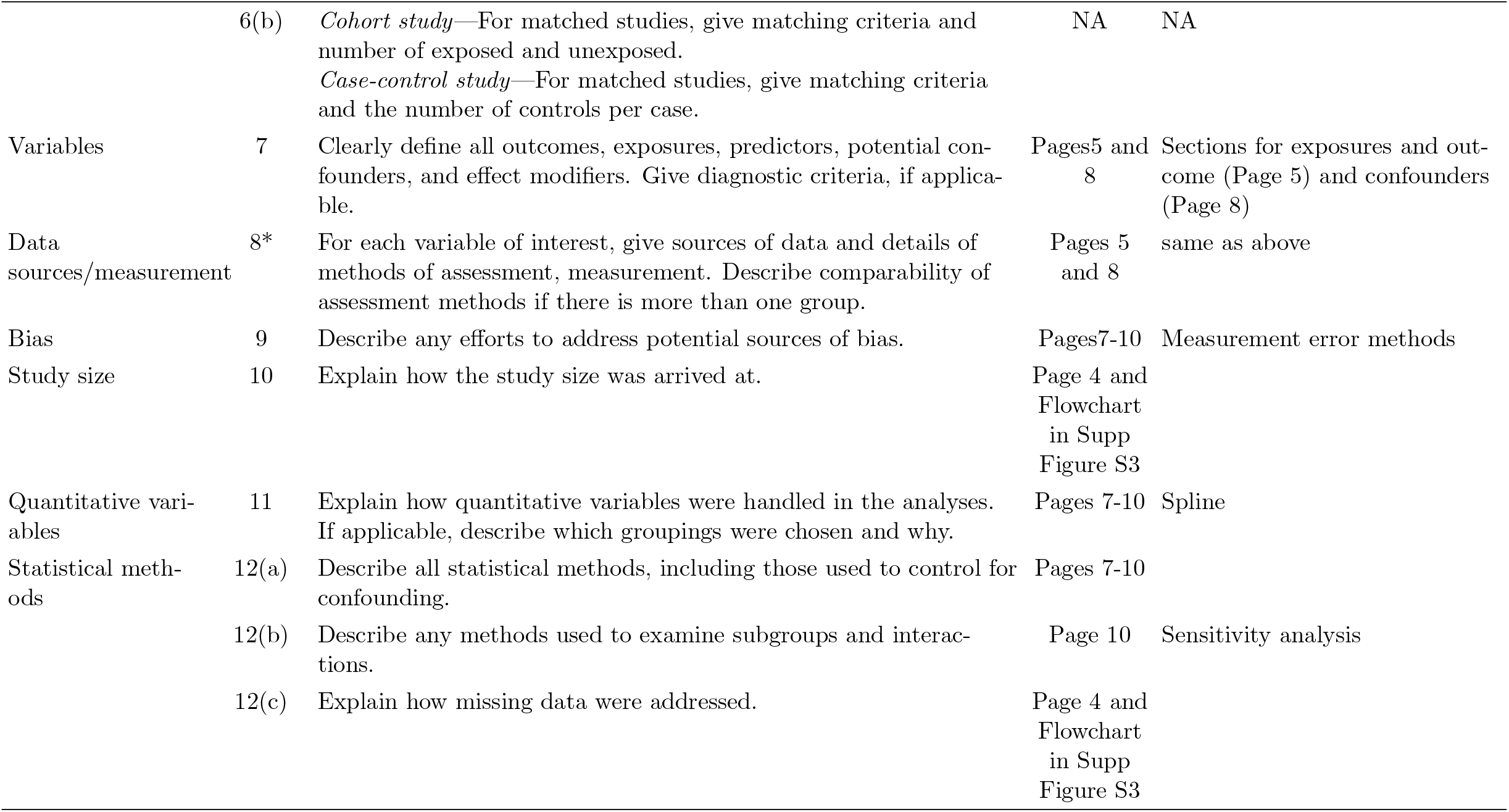
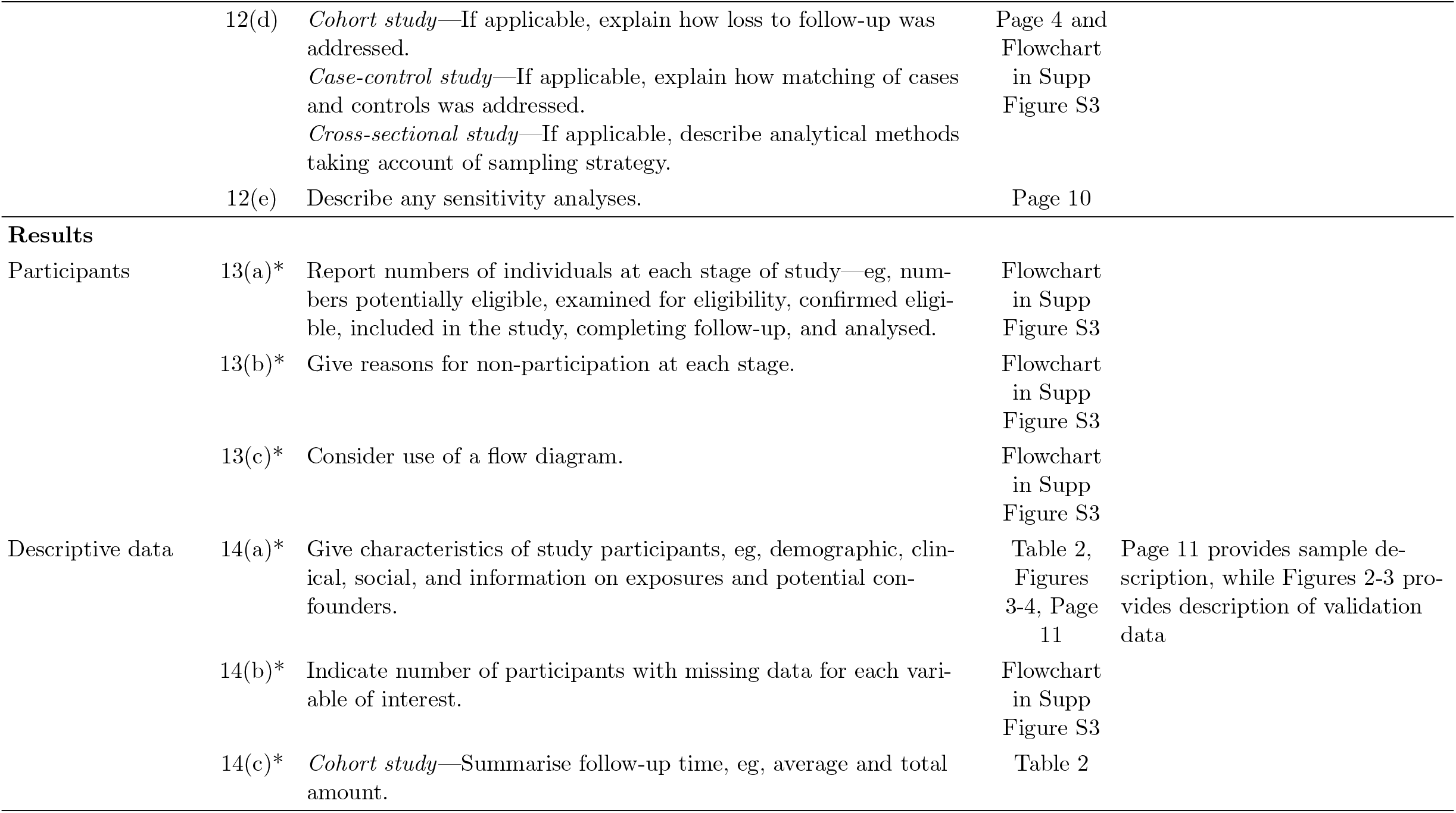
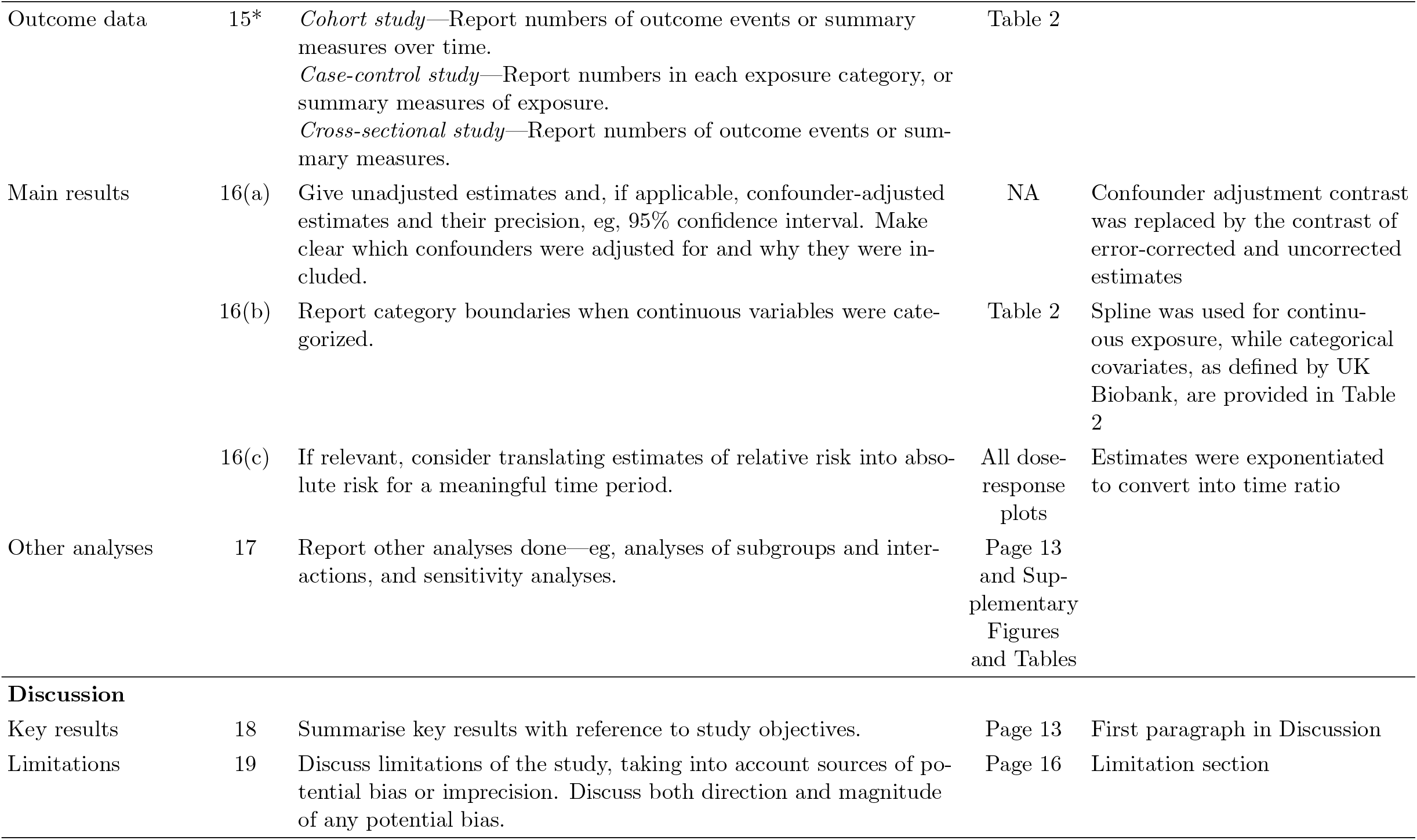
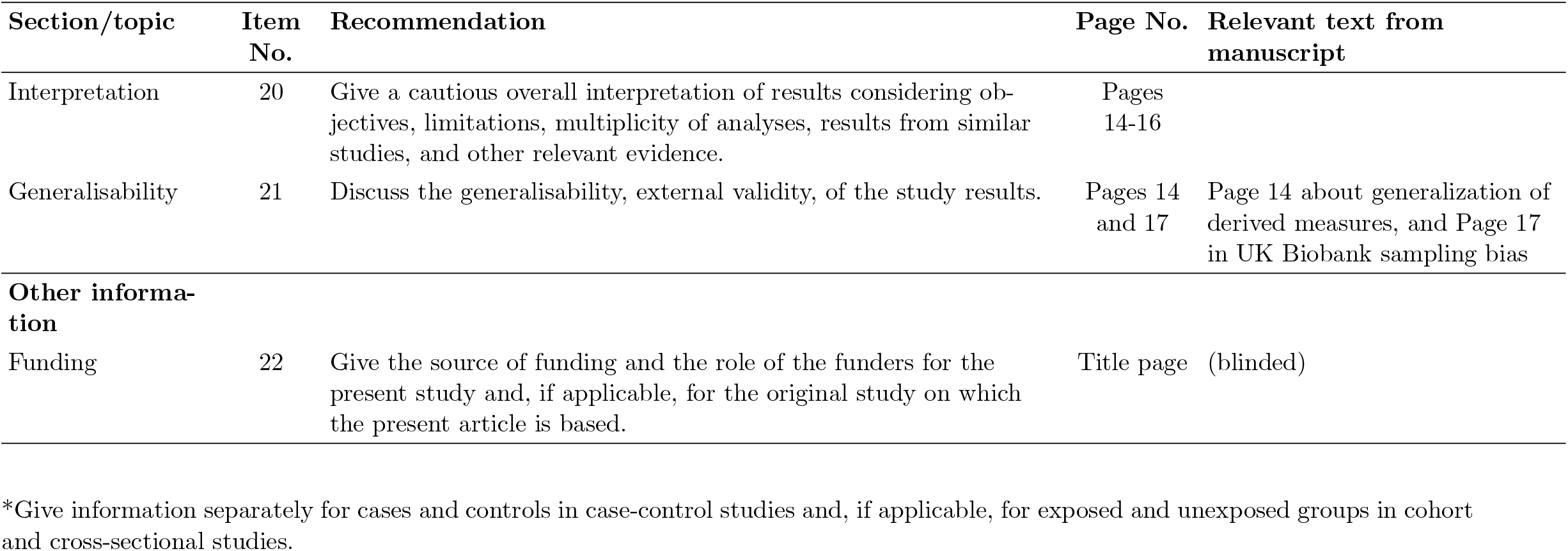

| Section/topic | Item No. | Recommendation | Page No. | Relevant text from manuscript |
| --- | --- | --- | --- | --- |
| **Title and abstract** | 1(a) | Indicate the study's design with a commonly used term in the title or the abstract. | Title page | Title contains survival analysis (cohort study) |
|  | 1(b) | Provide in the abstract an informative and balanced summary of what was done and what was found. | Page 1 | Model development, study design, exposure, outcome, simulation are added in abstract |
| **Introduction** |  |  |  |  |
| Background/rationale | 2 | Explain the scientific background and rationale for the investigation being reported. | Page 3 | Stated in Introduction |
| Objectives | 3 | State specific objectives, including any prespecified hypotheses. | Page 3 |  |
| **Methods** |  |  |  |  |
| Study design | 4 | Present key elements of study design early in the paper. | Page 3 | Stated in Introduction as survival analysis(cohort study), repeated in the beginning of Methods |
| Setting | 5 | Describe the setting, locations, and relevant dates, including periods of recruitment, exposure, follow-up, and data collection. | Page 4 | Data and population section |
| Participants | 6(a) | *Cohort study*—Give the eligibility criteria, and the sources and methods of selection of participants. Describe methods of follow-up.<br>*Case-control study*—Give the eligibility criteria, and the sources and methods of case ascertainment and control selection. Give the rationale for the choice of cases and controls.<br>*Cross-sectional study*—Give the eligibility criteria, and the sources and methods of selection of participants. | Page 4 | Data and population section and Flowchart in Supp Figure S3 |

Appendix 2. STROBE Checklist, continued
| Section/topic | Item No. | Recommendation | Page No. | Relevant text from manuscript |
| --- | --- | --- | --- | --- |
|  | 6(b) | *Cohort study*—For matched studies, give matching criteria and number of exposed and unexposed.<br>*Case-control study*—For matched studies, give matching criteria and the number of controls per case. | NA | NA |
| Variables | 7 | Clearly define all outcomes, exposures, predictors, potential confounders, and effect modifiers. Give diagnostic criteria, if applicable. | Pages 5 and 8 | Sections for exposures and outcome (Page 5) and confounders (Page 8) |
| Data sources/measurement | 8* | For each variable of interest, give sources of data and details of methods of assessment, measurement. Describe comparability of assessment methods if there is more than one group. | Pages 5 and 8 | same as above |
| Bias | 9 | Describe any efforts to address potential sources of bias. | Pages 7-10 | Measurement error methods |
| Study size | 10 | Explain how the study size was arrived at. | Page 4 and Flowchart in Supp Figure S3 |  |
| Quantitative variables | 11 | Explain how quantitative variables were handled in the analyses. If applicable, describe which groupings were chosen and why. | Pages 7-10 | Spline |
| Statistical methods | 12(a) | Describe all statistical methods, including those used to control for confounding. | Pages 7-10 |  |
|  | 12(b) | Describe any methods used to examine subgroups and interactions. | Page 10 | Sensitivity analysis |
|  | 12(c) | Explain how missing data were addressed. | Page 4 and Flowchart in Supp Figure S3 |  |
|  | 12(d) | *Cohort study*—If applicable, explain how loss to follow-up was addressed.<br>*Case-control study*—If applicable, explain how matching of cases and controls was addressed.<br>*Cross-sectional study*—If applicable, describe analytical methods taking account of sampling strategy. | Page 4 and Flowchart in Supp Figure S3 |  |
|  | 12(e) | Describe any sensitivity analyses. | Page 10 |  |
| **Results** |  |  |  |  |
| Participants | 13(a)* | Report numbers of individuals at each stage of study—eg, numbers potentially eligible, examined for eligibility, confirmed eligible, included in the study, completing follow-up, and analysed. | Flowchart in Supp Figure S3 |  |
|  | 13(b)* | Give reasons for non-participation at each stage. | Flowchart in Supp Figure S3 |  |
|  | 13(c)* | Consider use of a flow diagram. | Flowchart in Supp Figure S3 |  |
| Descriptive data | 14(a)* | Give characteristics of study participants, eg, demographic, clinical, social, and information on exposures and potential confounders. | Table 2, Figures 3-4, Page 11 | Page 11 provides sample description, while Figures 2-3 provides description of validation data |
|  | 14(b)* | Indicate number of participants with missing data for each variable of interest. | Flowchart in Supp Figure S3 |  |
|  | 14(c)* | *Cohort study*—Summarise follow-up time, eg, average and total amount. | Table 2 |  |

Appendix 2. STROBE Checklist, continued
| Section/topic | Item No. | Recommendation | Page No. | Relevant text from manuscript |
| --- | --- | --- | --- | --- |
| Outcome data | 15* | *Cohort study*—Report numbers of outcome events or summary measures over time.<br>*Case-control study*—Report numbers in each exposure category, or summary measures of exposure.<br>*Cross-sectional study*—Report numbers of outcome events or summary measures. | Table 2 |  |
| Main results | 16(a) | Give unadjusted estimates and, if applicable, confounder-adjusted estimates and their precision, eg, 95% confidence interval. Make clear which confounders were adjusted for and why they were included. | NA | Confounder adjustment contrast was replaced by the contrast of error-corrected and uncorrected estimates |
|  | 16(b) | Report category boundaries when continuous variables were categorized. | Table 2 | Spline was used for continuous exposure, while categorical covariates, as defined by UK Biobank, are provided in Table 2 |
|  | 16(c) | If relevant, consider translating estimates of relative risk into absolute risk for a meaningful time period. | All dose-response plots | Estimates were exponentiated to convert into time ratio |
| Other analyses | 17 | Report other analyses done—eg, analyses of subgroups and interactions, and sensitivity analyses. | Page 13 and Supplementary Figures and Tables |  |
| **Discussion** |  |  |  |  |
| Key results | 18 | Summarise key results with reference to study objectives. | Page 13 | First paragraph in Discussion |
| Limitations | 19 | Discuss limitations of the study, taking into account sources of potential bias or imprecision. Discuss both direction and magnitude of any potential bias. | Page 16 | Limitation section |
| Interpretation | 20 | Give a cautious overall interpretation of results considering objectives, limitations, multiplicity of analyses, results from similar studies, and other relevant evidence. | Pages 14-16 |  |
| Generalisability | 21 | Discuss the generalisability, external validity, of the study results. | Pages 14 and 17 | Page 14 about generalization of derived measures, and Page 17 in UK Biobank sampling bias |
| **Other information** |  |  |  |  |
| Funding | 22 | Give the source of funding and the role of the funders for the present study and, if applicable, for the original study on which the present article is based. | Title page | (blinded) |

\*Give information separately for cases and controls in case-control studies and, if applicable, for exposed and unexposed groups in cohort and cross-sectional studies.

## Notes

**Conflict of interest** The authors declare no conflicts of interest.

### Competing Interest Statement

The authors have declared no competing interest.

### Author Declarations

The Research Ethics Board of the Faculty of Medicine and Health Sciences of McGill University gave ethical approval for this work.

